# Harmonising Structural Brain MRI from Multiple Sites with Limited Sample Sizes

**DOI:** 10.64898/2026.04.21.26351106

**Authors:** Gaurav Bhalerao, Pawel Markiewicz, Jacob Turnbull, David L. Thomas, Enrico De Vita, Laura Parkes, Gerard Thompson, Jane MacKewn, Georgios Krokos, Catriona Wimberley, William Hallett, Li Su, Paresh Malhotra, Nigel Hoggard, John-Paul Taylor, David Brooks, Craig Ritchie, Joanna Wardlaw, Paul Matthews, Franklin Aigbirho, John O’Brien, Alexander Hammers, Karl Herholz, Frederik Barkhof, Karla Miller, Julian Matthews, Stephen Smith, Ludovica Griffanti

## Abstract

Harmonisation is widely used to mitigate site- and scanner-related batch variability in multisite neuroimaging studies and is particularly critical in longitudinal clinical trials, where detection of subtle biological or treatment-related changes depends on reliable measurement across scanners and timepoints. However, the effectiveness of harmonisation in small, heterogeneous clinical datasets remains insufficiently understood, particularly in relation to subject-level variability and consistency across acquisition settings, and its impact on both removal of technical variability and preservation of biological variation in pooled multisite analyses. We systematically evaluated a range of image-based and statistical harmonisation methods using a clinically realistic multisite, multiscanner structural T1-weighted (T1w) MRI test-retest dataset comprising three controlled acquisition scenarios: repeatability, intra-scanner reproducibility and inter-scanner reproducibility. Methods were applied under different batch specifications (site, scanner, or both) and performance was assessed within each scenario and in pooled data using a multi-metric framework capturing both technical and biological variability in volumetric imaging-derived phenotypes (IDPs) relevant to aging and dementia research. Across IDPs, before harmonisation variability was lowest in the repeatability scenario (median variability=0.6 to 2.7%, rank consistency ρ ≥0.9), with modest increases under intra-scanner reproducibility (0.5 to 3.2%, ρ=0.5 to 1.0) and substantially greater variability under inter-scanner reproducibility conditions (1.7 to 19.2%, ρ =−0.1 to 0.9). These results offer important information to consider for multisite study design, including sample size calculation in clinical trials. Harmonisation performance was strongly context dependent, with clearer benefits emerged in inter-scanner scenarios where both variability reduction and improvements in subject-level consistency were observed. In pooled data, approaches that explicitly modelled *site* as batch and accounted for repeated-measure structure showed greater consistency across IDPs in batch effect mitigation and more accurately reflected underlying biological variation. Our evaluation metrics enabled disentangling the removal of global batch effect while highlighting residual variability at the phenotype-specific or multivariate levels. These findings demonstrate that harmonisation cannot be treated as a one-size-fits-all solution and must be interpreted relative to the acquisition context, dataset structure, and downstream analytic goals. Multi-metric evaluation under realistic clinical constraints is essential to support reliable and translatable neuroimaging inference by ensuring appropriate correction of batch effects while preserving longitudinal biological signals and sensitivity to clinically meaningful change in multisite studies.

## 1. Introduction

Assessing and reducing site- and scanner-related batch variability is essential in multisite clinical MRI studies, where imaging data support clinical trial design, biomarker identification, and treatment evaluation in disease modifying therapies. Such variability can obscure true biological differences and undermine the reliability of longitudinal or cross-site comparisons. This has direct implications for clinical trials, as increased measurement variability can reduce statistical power, inflate required sample sizes and hinder the detection of treatment effects. Importantly, the magnitude of this technical variability is not uniform, but can differ substantially depending on whether repeated scans are acquired on the same scanner, across sites using similar hardware, or across different scanner manufacturers. These considerations are especially important in aging and dementia trials, which involve older participants and may be affected by ongoing age-related atrophy over time. Preserving this biological variability while minimising technical variability is crucial for detecting treatment effects and avoiding confounding of outcomes (Wittens et al., 2021). Despite advances in harmonisation methods aimed at addressing batch effects in neuroimaging datasets, most have been developed and evaluated using large, relatively homogeneous research cohorts (Marzi et al., 2024). In contrast, clinical trial-oriented datasets are typically smaller and more heterogeneous in their scanner and acquisition characteristics and include participants with diverse clinical profiles. This combination of technical complexity and biological heterogeneity makes harmonisation both indispensable and intrinsically more challenging in such settings. Assessing the extent of batch variability (site, scanner or acquisition related) and determining how effectively it can be reduced while preserving true biological effects, is therefore a crucial step in neuroimaging analyses. An important open question is whether methods that perform well in large, well-controlled research cohorts generalise to the smaller, more heterogeneous clinical datasets encountered in practice.

Harmonisation can be applied at different stages of the workflow from acquisition and preprocessing through to outcome metrics (also known as imaging-derived phenotypes or IDPs). Post-acquisition approaches are particularly common, as they offer a practical means of reducing site and scanner variability in datasets where acquisition was not standardised. However, even when protocols are nominally matched across sites, inherent differences in scanner hardware and vendor-specific sequence implementations can introduce systematic biases in the images, further motivating the need for post-acquisition harmonisation. Previous works have focused on applying such approaches at the image, analysis and IDP level (Bayer et al., 2022; Hu et al., 2023). For instance, (Bordin et al., 2021) identified the image pre-processing strategies to maximise the consistency across Whitehall II (*N* = 774) and UK Biobank (UKB) datasets (*N* = 2.2k) for the harmonisation of white matter hyperintensities. Similarly, (Y. Li et al., 2021) investigated the impact of preprocessing methods (such as bias field correction and image resampling) and different harmonisation methods in MRI-derived metrics (*N* = 28). Image-based harmonisation methods typically aim to reduce scanner-related variability by adjusting image characteristics such as intensity and contrast. Some examples include intensity normalisation (34 scans; (Wrobel et al., 2020)) and contrast harmonisation using a U-Net-based deep learning model (24 scans; (Dewey et al., 2019)). Recent developments have scaled to larger datasets such as the SynthSR framework (>10k scans; (Iglesias et al., 2023)) and other related deep learning approaches for structural MRI scans (Abbasi et al., 2024). For instance, a domain-adaptation model was trained on ∼6k samples (Dinsdale et al., 2021), and a structure-preserving embedding method for multiscanner harmonisation was proposed using similar deep learning strategy (Torbati et al., 2023). At the IDPs level, statistical methods are commonly used, with ComBat (N >=200; (Fortin et al., 2017, 2018)) and its extensions e.g., CovBat (*N* = 505, 65 sites; (Chen et al., 2022)), ComBat-GAM (*N* = 10k, 18 sites; (Pomponio et al., 2020)), and longitudinal ComBat (*N* = 663, 58 sites; (Beer et al., 2020)) among the most popular methods. ComBat-like approaches estimate batch effects using an empirical Bayes (EB) framework that require specifying the batch variable and assumes adequate sample sizes within each batch. While these conditions are not always met in practice particularly in datasets with anonymized or small or uneven batch sizes, the extent to which ComBat remains effective under such constraints warrants systematic investigation (Orlhac et al., 2022; Parekh et al., 2022). An alternative approach that addresses this limitation is using continuous measures of image quality (image quality metrics or IQMs) as proxy for batch variable, since IQMs capture underlying properties such as signal-to-noise ratio, contrast, and other acquisition-dependent characteristics that vary systematically across batches. IQM-based harmonisation methods are conceptually appealing for smaller datasets, where batch identifiers are unknown or may insufficiently capture batch variability; however, their evaluation to date has largely focused on large samples. For instance, a method proposed by (Garcia-Dias et al., 2020) used image quality metrics (IQMs) from images to predict the ComBat-harmonised brain volumes (*N* = ∼18k scans, 89 scanners) and trained a machine learning model to harmonise scans from unseen sites (N = 454, 14 scanners). A further challenge for IQM-based approaches is that some IQMs may correlate with biological or the outcome metrics, raising the risk that biological signal could be inadvertently removed. More recently (Prevot et al., 2025) replaced batch identifiers with IQMs and used a Bayesian regression trees framework to flexibly model relationships between IQMs and IDPs, thereby harmonising a cross-sectional dataset of ∼5k participants. Taken together, while image-based, statistical, and IQM-driven harmonisation methods have shown promise in large, well-controlled research cohorts, their performance in clinically important settings characterised by small sample sizes and heterogeneous batches remains largely unexplored. Critically, it is unclear how the widely used ComBat-based approaches behave under such constraints, whether methods that do not rely on explicit batch variables (such as IQM-based harmonisation) can offer advantages in repeated small sample size study designs, and to what extent image-based approaches provide robust and practical solutions for addressing variability in the outcome metrics from multisite clinical datasets.

In previous studies, harmonisation has been evaluated using a variety of strategies, often tailored to the dataset context and the goals of the chosen method (Hu et al., 2023), making harmonisation approaches difficult to compare. ComBat approaches evaluated whether the additive (mean shift) and multiplicative (scaling) batch effects were removed upon harmonisation (Beer et al., 2020; Fortin et al., 2017, 2018). (Pomponio et al., 2020) evaluated the ComBat-GAM approach by developing brain age models before and after harmonisation using regional brain volumes. (Garcia-Dias et al., 2020) evaluated the performance of their machine learning model on an external validation set using Kolmogorov-Smirnov test and coefficient of variation of brain volumes. Other studies have used other evaluation metrics such as Cohen’s d (Maikusa et al., 2021), Intra-class correlation (Gebre et al., 2023; Lu et al., 2025), preservation of biological covariates (Sun et al., 2022). However, existing works often emphasise specific evaluation metrics to characterise the harmonisation performance, and many do not jointly quantify reductions in technical variability and the preservation of meaningful biological signal. This highlights the need for more systematic assessments using different evaluation metrics,.

To address these gaps, we utilised the Dementias Platform UK (DPUK) PET-MR Harmonisation dataset, a multicentre initiative explicitly designed with clinical trial applications in mind (Pawel J. Markiewicz et al., 2026). Conducted in healthy elderly participants, it established a network of eight PET-MR scanners across the UK to develop harmonised scanning protocols and quantify both within-site repeatability and cross-site reproducibility, providing a shared resource to support future multicentre studies in ageing and dementia. Its modest sample sizes and substantial multisite variability make it a valuable testbed for assessing harmonisation performance under conditions more typical of clinical studies than of large research cohorts. The data was acquired into three experimental groups – repeatability (same scanner), intra-scanner (same scanner type, different site), and inter-scanner (different scanner type) acquisition setting, yielding heterogeneous data and unbalanced batches that introduce varying levels of site and scanner effects. We use this dataset as a challenging benchmark to evaluate how existing harmonisation approaches behave when sample sizes are limited and batch structure is complex. Within this setting, we systematically assessed both image-based (histogram matching, SynthSR) and statistical harmonisation methods (ComBat-like and IQM-based) using a comprehensive evaluation framework that quantifies technical and biological variability in clinically relevant volumetric IDPs across multiple levels. To reflect uncertainty in batch definition, each method was evaluated under alternative batch definitions capturing scanner- or site-related effects. By examining harmonisation effects across distinct experimental conditions as well as in pooled analyses, our goal is to characterise method-specific trade-offs in small, heterogeneous cohorts and to clarify the extent to which current harmonisation strategies can support reliable and clinically meaningful inference in multisite neuroimaging.

## 2. Methods

### 2.1 Dataset

This multisite harmonisation and test-retest study was conducted across all eight DPUK sites equipped with hybrid PET/MR scanners. Two scanner models were used: three Siemens Biograph mMR systems (Delso et al., 2011) and five General Electric (GE) Signa PET/MR systems (Grant et al., 2016). Healthy volunteers were recruited at each site and underwent two PET/MR scans with a standardised protocol of MRI sequences relevant to dementia research. Each participant was scanned at the recruiting site and the retest scan was then assigned by group randomisation to one of the three conditions: (1) same -scanner repeatability, in which both scans were performed on the same scanner; (2) intra-scanner reproducibility, in which the retest scan performed at a different site but on the same scanner model; or (3) inter-scanner reproducibility, in which the retest scan was performed at a different site using a different scanner model. Ethical approval for the study was granted by the appropriate UK Research Ethics Committee (North West Greater Manchester East, 18/NW/0102). Participants were recruited either through the Join Dementia Research platform (https://www.joindementiaresearch.nihr.ac.uk/) or through a letter of approach at participating sites. Written informed consent was obtained from all participants prior to enrolment. The study was performed in accordance with the ethical standards as laid down in the 1964 Declaration of Helsinki and its later amendments. More details on the study protocol and ethical approval are also reported elsewhere (Pawel J. Markiewicz et al., 2026). The MRI acquisition includes multiple modalities, including structural, functional, and diffusion imaging; however, in this study we focused on T1-weighted (T1w) MRI, as it provides structurally rich, clinically interpretable images and supports well-established volumetric markers of ageing and dementia. T1-weighted acquisitions were performed using vendor-specific T1w sequence implementations, with matched spatial resolution and inversion time across scanners, while other acquisition parameters differed (**Table 1**). We used T1w brain MRI scans from 34 healthy elderly participants (15 repeat-scanner group, 10 intra-scanner group, 12 inter-scanner group, two timepoints per participant acquired within approximately 1 year; age range 65–83 years; See **Table 2** for more details).

**Table 1.** Key acquisition parameters for T1w scans.

| Parameter | Siemens Scanner (N = 34) | GE Scanner (N = 40) |
| --- | --- | --- |
| Manufacturer model | Biograph mMR | SIGNA PET/MR |
| Field Strength (T) | 3T | 3T |
| TE (ms) | 2.96 | 2.996 |
| TI (ms) | 900 | 900 |
| Flip angle (degrees) | 9 | 8 |
| Voxel resolution (mm) | 1x1x1 | 1x1x1 |
| Image size | 208x240x256 | 164x256x256 |
| Acquisition time (mm:ss) | 05:12 | 05:29 |

**Table 2.** Dataset counts and age (mean, standard deviation) for each test-retest category.

| Characteristics | Repeatability | Reproducibility<br>Intra-scanner- model | Reproducibility<br>Inter-scanner |
| --- | --- | --- | --- |
| Total counts | 15 | 10 | 12 |
| GE scanner counts | 8 | 6 | 8 |
| Siemens scanner counts | 7 | 4 | 4 |
| Baseline age | 71.7 ± 6.0 | 72.9 ± 5.5 | 72.6 ± 4.5 |

### 2.2 Image processing

The DICOM (Digital Imaging and Communications in Medicine) images were converted to NIfTI (Neuroimaging Informatics Technology Initiative) format using the dcm2niix tool (v1.0.20230411) (X. Li et al., 2016). All images were visually checked to ensure that all the slices were acquired, and the entire head was covered. Qualitative visual inspection indicated consistent Grey–White contrast across timepoints for repeat- and intra-scanner acquisitions, with no obvious changes in overall image appearance. In contrast, inter-scanner comparisons showed more noticeable differences in contrast and intensity characteristics (**Figure 1**). Images were also inspected to identify any low-quality scans to get an indication of subject or scanner-related artefacts. After the visual checks, all T1w scans were processed using an adapted version of the UKB image processing pipeline, to extract volumetric image derived phenotypes (IDPs) (Alfaro-Almagro et al., 2018; Bhalerao, Gillis, Alfaro-Almagro, et al., 2025). The pipeline primarily uses FSL tools (Smith et al., 2004) such as brain extraction (FSL-BET), bias correction and tissue segmentation (FSL-FAST), subcortical segmentation (FSL-FIRST) and extraction of normalised (corrected for head size) brain and tissue volumes (FSL-SIENAX) to generate volumetric measures from T1w scans. Although the pipeline extracts numerous IDPs from T1w scans, our analyses focused on a subset most pertinent to clinical and diagnostic applications in aging and dementia: total brain volume, grey matter (GM), peripheral GM, white matter (WM), and cerebrospinal fluid (CSF) volumes, and left and right hippocampus volumes.

**Figure 1.**
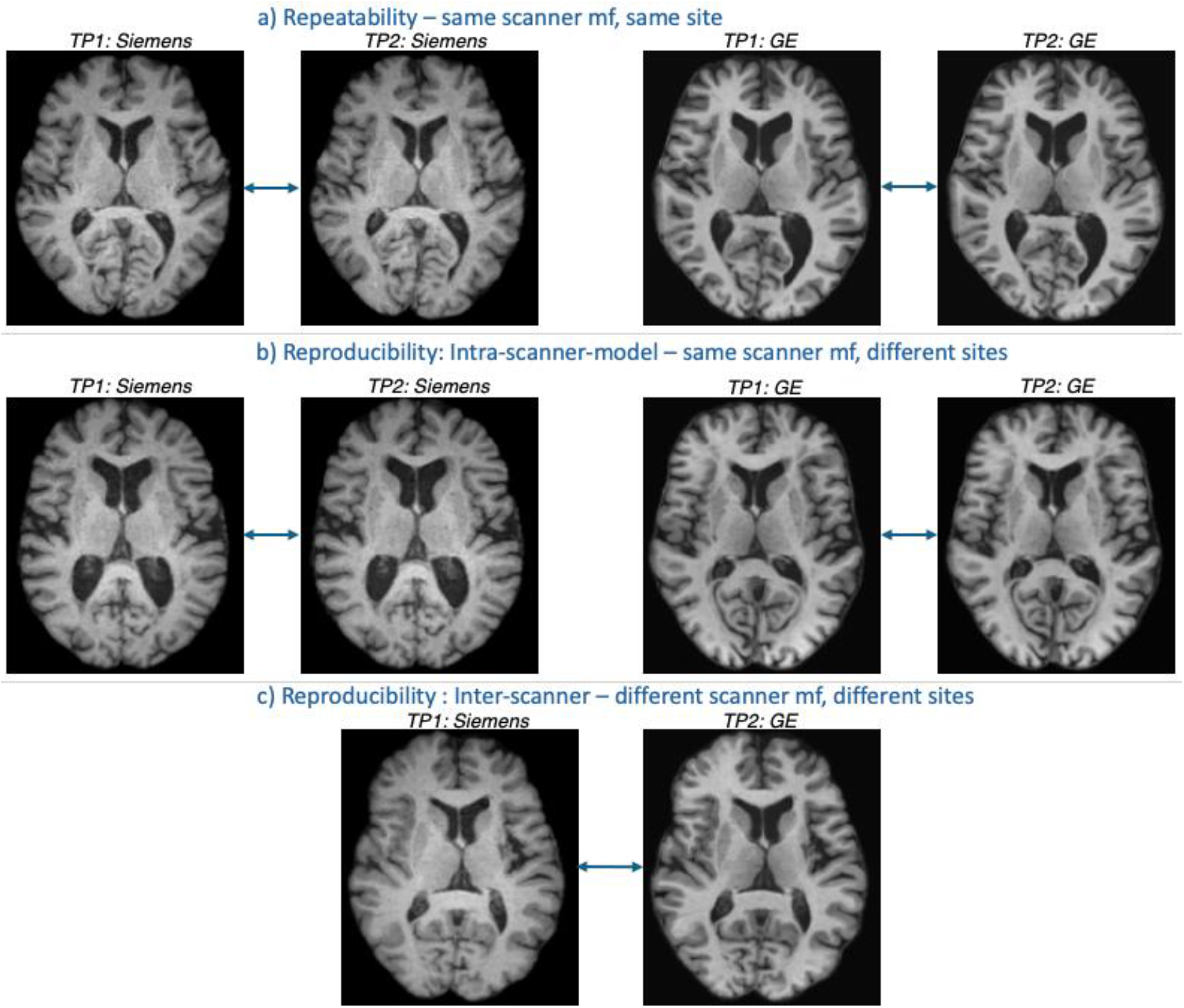
Representative T1-weighted images illustrating image similarity and contrast differences across three test-retest categories: a) repeatability: same subject scanned twice on the same scanner manufacturer (mf) either Siemens or GE; b) intra-scanner-model reproducibility: same subject scanned on scanners from the same mf (either Siemens or GE) at different sites; c) inter-scanner reproducibility: same subject scanned on scanners from different mf at different sites. Visual inspection shows consistent grey-white matter contrast for repeat- and intra-scanner acquisitions, with more noticeable contrast differences in the inter-scanner comparison.

### 2.3 Assessing variability in raw IDPs

We assessed scanner-related variability in raw or original IDPs using two complementary metrics, evaluated separately across three test-retest categories (repeat-scanner, intra-scanner, and inter-scanner). This stratified analysis isolates the effects of increasing scanner heterogeneity on within-subject variability and subject-wise correspondence, providing a measure of subject-level stability and diagnostic basis for the subsequent application of harmonisation methods.

#### 2.3.1 Within-subject variability

To quantify within-subject variability, for each subject, we computed the Relative Percent Difference (RPD) in IDPs between the two timepoints. Lower RPD values indicated lower within-subject variability across timepoints reflecting consistency in the measurements and reduced scanner-related (non-biological) variability. RPD was calculated as the absolute difference between the two timepoints divided by their mean, expressed as a percentage for each IDP and each subject (Eq. 1):

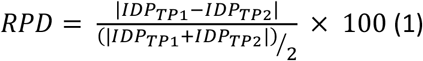

#### 2.3.2 Subject order consistency

Because these test-retest scans were acquired over a relatively short interval (within one year for most subjects), large changes in the relative ordering of subjects according to IDP values between the two timepoints are unlikely, even in the presence of ongoing biological change. Accordingly, preservation of subject ranking across timepoints provides a useful measure of within-subject consistency. To quantify this, we computed the Spearman rank correlation (ρ) between timepoints for each IDP (Warrington et al., 2023). Spearman correlation provides a non-parametric measure of monotonic association and is robust to non-normality and outliers, making it well suited for assessing rank consistency across timepoints. Higher correlation values indicate higher preservation of subject ranking across timepoints, reflecting greater within-subject consistency. Statistical inference was performed using a permutation test assessing the null hypothesis of no within-subject association (i.e., random subject ordering across timepoints). For each IDP, subject labels at the second timepoint were randomly permuted (n = 10,000 permutations) while the first timepoint were held fixed, generating a null distribution of Spearman *p* values under random pairing. Because the a priori hypothesis was positive association between timepoints, inference used a one-sided (upper-tail) test. The permutation p-value was computed as (Eq. 2):

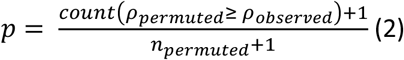

Where *ρ*_*observed*_ is observed Spearman correlation and *ρ*_*permuted*_ denotes permuted correlations. A significant result (p<0.05) indicates that the observed correlation is greater than expected under random subject ordering, consistent with preservation of subject-specific relative ranking over time.

### 2.4 Harmonisation methods

Throughout this work, we use the term *batch* to denote the factor used to model non-biological, technical variability in the scans or derived IDPs. In the present dataset, batch effects may arise at the level of scanner manufacturer, site, or a combination of the two. Note that each site operates a single scanner manufacturer (either GE or Siemens), such that scanner and site effects are not independently identifiable. To account for scanner- and site-related batch variability, several image-based and statistical harmonisation methods were applied (see **Table 3** for a summary). Harmonisation was performed on data pooled across three (repeat-scanner, intra-scanner, and inter-scanner) test-retest categories. This pooling reflects typical real-world multisite analysis scenarios, in which harmonisation is applied to combined datasets rather than stratified subsets. Pooling also increased the effective sample size available for estimating scanner- and site-level effects, as scanners and sites were shared across test-retest categories.

**Table 3.** Harmonisation methods used in this study. For statistical harmonisation methods, batch effects were modelled as either scanner, site or mixed-effects formulations.

| Method | Data type | Model class | Mean adjustment | Variance adjustment | Covariance adjustment | Random effects | Parameters estimated | Batch specification |
| --- | --- | --- | --- | --- | --- | --- | --- | --- |
| Histogram matching | Image | CDF-based intensity mapping | - | - | - | - | Non-parametric | Within-subject intensity |
| SynthSR | Image | Pre-trained CNN | - | - | - | - | Pre-trained model | Within-subject contrast |
| Linear regression (LR) | IDP | Additive fixed-effects model | Yes | No | No | No | Batch, Age, Timepoint | Scanner; Site |
| Linear mixed effects (LME) | IDP | Hierarchical mean model | Yes | No | No | Subject, Site | Fixed effect of Batch, Age, Timepoint; Random effect of Subject | Scanner; Site; Scanner fixed, Site random |
| ComBat | IDP | EB (additive + multiplicative ) | Yes | Yes | No | No | Age, Timepoint, additive, multiplicative batch effects | Scanner; Site |
| Longitudinal ComBat | IDP | EB + mixed-effects extension | Yes | Yes | No | Subject, Site | ComBat parameters, Random effect of Subjects | Scanner; Site; Scanner fixed, Site random |
| CovBat | IDP | Multivariate EB + PCA covariance harmonisation | Yes | Yes | Yes | No | ComBat parameters, PC score adjustments | Scanner; Site |
| IQM-based | IDP | Mixed-effects model with IQMs | Yes | No | No | Subject | Fixed effects of IQMs, Age, Timepoint; Random effect of Subjects | Scanner; Site |

#### 2.4.1 Image-based harmonisation methods

Two image-based harmonisation methods were applied on the paired scans from each subject to obtain the harmonised images. Image-based harmonisation methods below operate directly on image intensities and were included to assess whether batch differences can be mitigated prior to feature extraction. Histogram matching provides a simple, non-parametric intensity normalization baseline, while SynthSR uses deep learning–based synthesis to generate images with standardized contrast properties, offering robustness to scanner and protocol variability.

##### 1. Histogram matching (HistMatch)

For each pair of scans from the same subject, histogram matching was applied to compensate for batch effects by aligning the cumulative distribution functions of image intensities (Wrobel et al., 2020). All images were bias-corrected beforehand to remove low-frequency intensity inhomogeneities and help ensure that histogram alignment reflected true tissue contrast rather than scanner artifacts. For the repeated and intra-manufacturer groups, the first timepoint was used as the reference, whereas for the inter-manufacturer group, data from the Siemens scanner was used as the reference. The method was implemented using the ANTs toolkit (ImageMath function)(Avants BB et al., 2013).

##### 2. SynthSR

SynthSR is a convolutional neural network that converts scans of any orientation, resolution, or contrast into 1 mm isotropic MP-RAGE (Iglesias et al., 2023). Although our data were already acquired as 1 mm isotropic MP-RAGE, we selected SynthSR for its simplicity of implementation and its ability to handle clinical scans of varying contrast and resolution. In this context, our aim was to harmonise within-subject image contrasts while assuming preservation of the underlying anatomy. SynthSR was applied to all raw T1-weighted images using default settings in FreeSurfer 7.4.1 (Fischl, 2012).

The harmonised scans from histogram matching and SynthSR methods were then processed through the UKB pipeline to extract the IDPs. Intermediate processing steps (such as tissue segmentation quality) in the pipeline were visually checked to ensure accuracy.

#### 2.4.2 Statistical harmonisation methods

Various statistical methods were applied to the IDPs derived from raw T1-weighted images to produce harmonised IDPs. To account for non-biological variability at different levels, harmonisation methods were applied using three batch-variable formulations (where applicable). First, *scanner* manufacturer was modelled as a batch effect with two levels (GE vs Siemens), reflecting manufacturer-level differences observed in the test–retest analyses. Second, *site* was modelled as a batch effect with eight levels, capturing site-specific sources of variability that may persist within the same scanner manufacturer. Third, a mixed-effects formulation in which *scanner was modelled as a fixed batch effect and site as a random effect*, allowing site-level variability to be partially pooled across sites. This formulation is suitable in the present dataset since the number of sites is relatively large and per-site sample sizes are limited. Together, these formulations reflect different assumptions about the level at which non-biological variability operates in multisite data. The statistical methods described below were grouped into two broad categories: (1) cross-sectional harmonisation approaches that treat repeated observations as independent (linear regression, ComBat, CovBat); and (2) approaches that incorporate subject-level random effects to account for within-subject dependence (linear mixed-effects models, image quality metric–based, and Longitudinal ComBat). All analyses, unless specified otherwise, were performed in MATLAB R2024b (*MATLAB Version: 24.2.0.2863752 (R2024b)*, 2024).

##### 1. Linear regression (LR)

Batch effects were harmonised using a linear regression approach, included as a simple and widely used baseline method for adjusting IDPs for batch effects. This method provides a transparent reference point for evaluating the added benefit of more complex harmonisation models in the test– retest setting. For each IDP, we fitted a model of the form (Eq. 3),

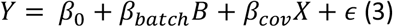

where B represents categorical batch variable and X represents biological covariates (age, timepoint). The estimated batch component *β*_*batch*_*B* was then subtracted, producing harmonised IDPs (*Y*^∗^ = *Y* − *β*_*batch*_*B*) while retaining variation due to biological covariates. The method was applied in two batch-variable formulations – 1) using scanner as batch effect (LR-Scanner), 2) site as batch effect (LR-Site).

##### 2. ComBat

Batch effects were further addressed using parametric ComBat, an EB method that models and removes additive and multiplicative batch effects while preserving biological covariates. ComBat was included due to its widespread adoption in neuroimaging and its ability to stabilise batch-effect estimates in multisite datasets with limited sample sizes (Fortin et al., 2018; Johnson et al., 2007). For each IDP, ComBat adjusts mean and variance across batches by modelling (Eq. 4),

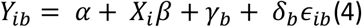

Where *Y*_*ib*_ is the IDP value for subject i in batch b; *α* is a global intercept; *X*_*i*_ contains biological covariates (age, timepoint); *β* are covariate coefficients; *γ*_*b*_ represents additive batch effects; *δ*_*b*_ represents multiplicative (variance-scaling) batch effects; and *ϵ*_*ib*_ ∼ 𝒩(0, *σ*^2^). Batch-specific parameters (*γ*_*b*_,*δ*_*b*_) are estimated using parametric EB, which shrinks batch estimates toward pooled values to improve stability when batch sample sizes are small.

After parameter estimation, harmonised IDPs are obtained by removing both additive and multiplicative batch effects while retaining biological covariate effects (Eq. 5):

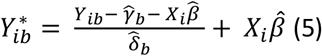

We ran two batch-variable formulations using ComBat: 1) scanner as batch variable (ComBat-Scanner) and 2) site as batch variable (ComBat-Site).

##### 3. CovBat

We applied CovBat (Chen et al., 2022), an extension of ComBat that adjusts not only the mean and variance but also the feature covariance structure across batches. CovBat was included to assess whether harmonisation of inter-feature covariance improves consistency of multivariate IDPs across sites in our dataset.

Unlike ComBat, which models each IDP independently, CovBat operates jointly on the vector of IDPs. Let *Y*_*i*_ ∈ ℝ^*q*^ denote the vector of q IDPs for subject i in batch b. After standard ComBat mean and variance adjustments, CovBat models residuals as:

For each IDP, the method models (Eq. 6),

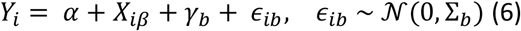

Where *α* is a global intercept; *X*_*i*_ contains biological covariates (age, timepoint); *β* are covariate coefficients; *γ*_*b*_ represents additive batch effects; Σ_*b*_ is the batch-specific covariance matrix across IDPs. Direct estimation of full qxq covariance matrices Σ_*b*_ would be unstable when q is large relative to batch sample size. CovBat addresses this by – 1) performing principal component analysis (PCA) on ComBat-adjusted residuals 2) harmonising batch effects in the leading PC scores and 3) reconstructing data in the origin feature space. Thus, covariance harmonisation is achieved in a reduced-dimensional representation rather than directly estimating and inverting high-dimensional covariance matrices. Parameter estimation includes mean and variance parameters per IDP (as in ComBat) and batch-specific adjustments of PC scores (for selected components). After estimation, harmonised IDPs are obtained by removing batch-related effects in both the marginal distributions (mean and variance) and in the leading covariance structure, while preserving covariate-related signal.

We ran two batch-variable formulations using CovBat: 1) scanner as batch effect (CovBat-Scanner) and 2) site as batch effect (CovBat-Site). CovBat was implemented using R (v4.4.2) (https://github.com/andy1764/CovBat_Harmonization) (R Core Team, 2022).

##### 4. Longitudinal ComBat (LongComBat)

To account for repeated measurements in our dataset (2 timepoints per subject), we applied a parametric version of longitudinal ComBat (Beer et al., 2020), an extension of ComBat that incorporates subject-level random effects. This method was included to explicitly model within-subject dependence while correcting additive and multiplicative batch effects, making it appropriate for test– retest and longitudinal acquisition designs. For each IDP (modelled independently), longitudinal ComBat assumes (Eq. 7):

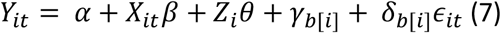

where *Y*_*it*_ is the IDP value for subject i at time t; *α* is a global intercept; *X*_*it*_ represents biological covariates (e.g., age, timepoint); *β* are fixed covariate effects; *Z*_*i*_*θ* captures the subject-specific random intercept (or slope) to model within-subject correlation; *γ*_*b*[*i*]_ and *δ*_*b*[*i*]_ are the additive and multiplicative batch effects, and *ϵ*_*it*_ ∼𝒩(0, *σ*^2^) is residual error. As in standard ComBat, batch parameters are estimated using EB shrinkage, but the model additionally partitions variance into between-subject and residual components to preserve within-subject structure. After model fitting, harmonised IDPs are obtained by removing estimated additive and multiplicative batch effects while retaining both fixed covariate effects and subject-specific random effects. Conceptually, this corresponds to (Eq. 8):

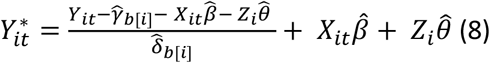

Thus, compared with ComBat, longitudinal ComBat additionally preserves subject-level variance components, preventing inflation or attenuation of within-subject correlations after harmonisation.

We ran three batch-variable formulations using longitudinal ComBat: 1) scanner as batch effect (LongComBat-Scanner), 2) site as batch effect (LongComBat-Site), 3) modelling scanner as a fixed batch effect and site as a random effect (LongComBat-ScannerFSiteR). Longitudinal ComBat was implemented using R (v4.4.2) package (R Core Team, 2022).

##### 5. Linear mixed effects (LME)

In contrast to the EB framework used in ComBat-based approaches, linear mixed-effects (LME) models estimate batch and biological effects simultaneously within a single hierarchical model. Batch effects are modelled as fixed and/or random effects, while subject-level random effects account for within-subject dependence across timepoints. For each IDP (modelled independently), we fitted LME models of the form (Eq. 9),

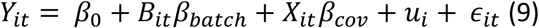

Where *Y*_*it*_ is the IDP for subject i at timepoint t; *β*_0_is the intercept; *B*_*it*_ encodes batch; *β*_*batch*_are batch-effect coefficients; *X*_*it*_ contains biological covariates (e.g., age, timepoint); *β*_*cov*_ are fixed covariate effects; 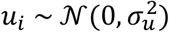) is a subject-specific random intercept capturing within-subject variability; *ϵ*_*it*_∼𝒩(0, *σ*^2^) is residual error. After fitting, harmonised IDPs are obtained by removing the estimated fixed batch effects (Eq. 10):

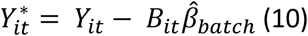

while retaining estimated biological effects and subject-specific random effects. When batch was modelled as a random effect, harmonisation corresponded to removing the estimated batch-specific random intercepts. We ran three batch-variable formulations using LME: 1) scanner as batch effect (LME-Scanner), 2) site as batch effect (LME-Site), 3) modelling scanner as a fixed effect and site as a random effect (LME-ScannerFSiteR).

##### 6. Image quality metrics (IQM)-based harmonisation (LME-IQM)

Building on previous works (Garcia-Dias et al., 2020; Prevot et al., 2025), and our previous study on using IQMs to design QC classifiers (Bhalerao et al., 2025), we extended the approach by implementing IQM-based harmonisation within a mixed effects framework. This approach was included to explicitly account for acquisition-related image quality variability that may confound batch effects and biological signal.

In this approach, we first extracted a set of quality metrics from each T1w scan using MRIQC (Esteban et al., 2017), CAT12 segmentation (Gaser et al., 2024), and DSMRI framework (Domain shift analyzer for MRI) (Kushol et al., 2023) tools. More details on IQMs from MRIQC and CAT12 can be found in (Bhalerao et al., 2025), with implementation details openly available (https://git.fmrib.ox.ac.uk/mcz502/qc-paper), while details on DSMRI implementation are available at (https://github.com/rkushol/DSMRI). In our previous work, we found that these tools capture overlapping as well as unique QC metrics, hence we used all metrics (total 117, see supplementary excel sheet – ‘*IQM-harmonisation_IQMlist’* for the list of IQMs) from these tools to capture as much information as possible. To reduce dimensionality and collinearity, IQMs were standardised and subjected to principal component analysis (PCA), retaining components explaining 95% of the total IQM variance.

IQM-based harmonisation was performed in two stages: 1) additive batch correction, 2) multiplicative batch correction. Because IQMs may correlate with biological variables of interest (e.g., age) and/or with valid signal of interest in the image-derived phenotypes (IDPs), we employed model-based filtering strategies to identify IQM components that predominantly captured technical variation but not biological variation. In the additive batch correction, for each IDP and each PCA-IQM component, mixed-effects models were fitted to identify components that were significantly associated with batch while not being associated with age or the IDPs (See *Appendix A* for full model specifications – Eq. (1), Eq. (2)). Only such components were retained for harmonisation.

First, additive batch effects were modelled by fitting an LME model of the form (Eq. 11):

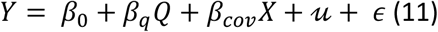

Where Y denotes the IDP, Q the selected PCA-IQM components, X biological covariates (age and timepoint), 𝓊 a subject-specific random intercept and *ϵ* residual error. Additive batch effect correction was applied by subtracting the estimated IQM-associated fixed-effect contribution *β*_*q*_*Q* from each observation, thereby removing QC-driven mean shifts while preserving biological effects.

The framework was subsequently extended to model multiplicative (scaling) batch effects. To identify IQM components capturing site-dependent scaling, a variance proxy was first derived from the log of squared residuals obtained from a mixed-effects model including biological covariates (age and timepoint) and subject-specific random effects (See Appendix A – Eq. (5), Eq. (6)). Each IQM component was then modelled as a function of batch, biological covariates (age and timepoint), and this variance proxy. IQM components were retained for multiplicative correction if they were significantly associated with batch (p < 0.05) but not significantly associated with residual variance proxy or age (*p* > 0.05), thereby targeting batch-related scaling while avoiding rescaling driven by subject-specific noise or test–retest variability (See Appendix A – Eq. (7), Eq. (8)). Multiplicative effects were evaluated in log-space on the additively corrected IDPs (Eq. 12).

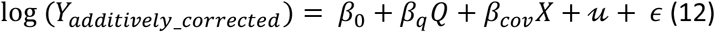

Multiplicative correction was applied only when inclusion of the selected IQM components significantly improved model fit based on likelihood-ratio model comparison (See Appendix – Eq. (10), (11)); otherwise, no multiplicative scaling was performed.

We applied this method with two batch-variable formulation, one with scanner as batch variable (LME-IQM-Scanner) and other with site as batch (LME-IQM-Site).

###### Exploratory identification of dominant IQMs

In an exploratory analysis, we sought to identify the IQMs that were dominant in driving batch-effect correction. This analysis was intended to provide insight into the structure of QC contributions underlying different batch formulations and correction strategies.

To summarise our method above, for each batch formulation, additive and multiplicative correction strategies were applied using distinct filtering criteria to select PCA–IQM components for each IDP. First, PCA components selected for additive and multiplicative correction were identified separately for each IDP. For each selected component, PCA loadings were examined and the five IQMs with the largest absolute loadings were identified as the dominant contributors to that component. This procedure yielded a set of IQMs associated with each IDP. Next, IQMs identified across all IDPs were aggregated to obtain the union of dominant IQMs for each correction strategy.

This analysis was performed independently for the two batch formulations (scanner as batch and site as batch). Unique and overlapping IQMs across these batch formulations were then identified to characterise QC metrics that were consistently important, as well as those that appeared specific to scanner- or site-related batch effects.

### 2.5 Harmonisation evaluation

Although harmonisation methods were applied using different batch-variable formulations (scanner, site, or scanner as a fixed effect with site as a random effect), all harmonisation outcomes were evaluated with respect to site-level effects. Site was selected as the evaluation batch variable because it represents the most granular source of non-biological variability in the dataset and subsumes scanner-related effects, given that each site operates a single scanner manufacturer. This strategy provides a consistent and conservative benchmark for comparing harmonisation methods and enables assessment of whether correction based on coarser batch definitions (e.g., scanner alone) is sufficient to remove residual site-level effects. We applied a comprehensive set of evaluation metrics to assess method performance before and after harmonisation (**Table 4** for the summary of these metrics). Two metrics (2.5.1, 2.5.2) were evaluated on each group (repeat-scanner, intra-scanner, inter-scanner) whereas model-based diagnostics (2.5.3 onwards) were performed on the pooled dataset from all three groups to obtain stable estimates across batches.

**Table 4.** Harmonisation evaluation framework for the test-retest dataset

| Variability | Procedure | Interpretation of effect before harmonisation | Expected effect after successful harmonisation | Interpretation of effect after harmonisation |
| --- | --- | --- | --- | --- |
| Additive effects* | Fit linear mixed-effects model per IDP with batch and biological covariates as fixed effect and subject as random effect. Compare full model (with batch) to reduced model (without batch) using Kenward-Roger F-test. | Significant test statistic indicates presence of additive batch effect | Reduce | Reduced F-statistics and non-significant p-values indicate removal of additive batch effects |
| Multiplicative effects* | Fit linear mixed-effects model per IDP with batch as fixed effect and subject as random effect. Test homogeneity of variance for residuals across batches using Fligner-Killeen test. | Significant variance heterogeneity across batches indicates multiplicative batch effects. | Reduce | Reduced chi-squared statistic and non-significant p-values indicate removal of variance-related batch effects. |
| Site-pairwise batch effects | Fit linear mixed-effects model with IDP as response, batch and biological covariates as fixed effects, and subject as random effect. Conduct pairwise site comparisons using linear contrasts. | Significant pairwise differences indicate residual site-specific mean differences. | Reduce | Non-significant pairwise site differences indicate minimal residual batch effect. |
| Multivariate batch effect | Calculate distance between the reference and each batch using mean and covariance across IDPs | Large MD values indicate multivariate divergence of site distributions due to batch effects. | Reduce | Reduced MD values indicate better multivariate alignment across sites. |
| Subject order consistency | Calculate Spearman rank correlation between the timepoints for each IDP. | Low rank correlation indicates inconsistent subject ordering due to measurement noise or batch effects. | Preserve/Improve | Higher rank correlation indicates consistent subject order across the timepoints. |
| Within-subject variability | Calculate relative percentage difference for IDPs between the | High variability indicates poor test- | Reduce | Reduced variability indicates IDPs between paired scans are similar. |
|  | two timepoints for each subject. | retest reliability and measurement noise. |  |  |
| Between-subject variability | Fit mixed-effects model with IDP as response and subject as random intercept. Calculate the ICC from between-subject variance and residual variance. | Low ICC indicates dominance of residual or site-related variance over subject differences. | Preserve/Improve | Preserved/improved ICC indicates within-subject variability is reduced with respect to between-subject variability. |
| Biological variability | Fit linear mixed-effects model with IDP as response, age and timepoint as fixed effects, and subject as random intercept. Extract effect sizes, confidence intervals, and p-values. | Attenuated or inconsistent associations may reflect site-related confounding or measurement noise. | Preserve/Reveal true effects | Effect sizes and significance should be preserved, or true effects should be revealed post-harmonisation. |

\*Implemented in R version 4.4.2; other metrics implemented in MATLAB 2024b.

#### 2.5.1 Within-subject variability

To quantify within-subject variability (same as 2.3.1), we computed the relative percent difference for each subject and IDP between timepoints, both before and after harmonisation. Lower RPD values indicate reduced within-subject variability. RPD was calculated as the absolute difference between the two measurements divided by their mean, expressed as a percentage (See Eq. 1). For each test-retest category, median RPD values were computed across subjects for each harmonisation method. To statistically assess whether harmonisation altered within-subject variability, paired comparisons between raw and harmonised RPD values were performed using the Wilcoxon signed-rank test (two-sided, *α* =0.05). A significant decrease in RPD after harmonisation was interpreted as improved measurement consistency and reduced non-biological variability. Conversely, a significant increase in RPD indicated a deterioration in within-subject consistency following harmonisation.

#### 2.5.2 Subject order consistency

Given the short interval between acquisitions, any ongoing biological change is unlikely to result in large shifts in the relative ordering of subjects across IDPs over this period. Subject-order consistency across timepoints can therefore be used as a negative control for harmonisation-induced distortions and harmonisation should not disrupt subject ranking and may improve it by reducing measurement noise. To quantify preservation of subject ordering (same as 2.3.2), we computed the Spearman rank correlation between timepoints for each IDP, both before and after harmonisation (Warrington et al., 2023). An increase in correlation after harmonisation suggests that harmonisation reduced non-biological variability without distorting subject-specific differences. The correlation values were calculated and compared before and after harmonisation for each test-retest category to see how each harmonisation method impacted the rank consistency across the timepoints in each group.

#### 2.5.3 Additive and multiplicative batch effects

Because ComBat-based approaches explicitly model and correct additive (mean) and multiplicative (variance) batch effects, we evaluated these effects before and after harmonisation to assess whether such batch-related variability was successfully removed from the IDPs. We used the code provided in (Beer et al., 2020) to estimate the additive and multiplicative effects for each raw and harmonised IDP.

##### Additive batch effect

Additive batch effect was quantified for each IDP by fitting mixed-effects models with IDP as the dependent variable, biological covariates (age, timepoint) as fixed effects and subjects as random effect. A full model including site as batch effect was compared with a reduced model excluding batch using a Kenward–Roger F-test, yielding estimates of the significance of additive batch effects. This model comparison yields an omnibus test of whether site explains additional variance in the IDP beyond biological covariates before and after harmonisation. IDPs with *p* > 0.007 (Bonferroni corrected for 7 IDPs, 0.05/7=0.007) were considered to have no significant additive batch effects.

##### Multiplicative batch effect

Multiplicative batch effect was assessed for each IDP by fitting linear mixed-effects models with IDP as the dependent variable, site as batch effect, biological covariates as fixed effects and subject as random effect. Residuals from this full model were tested for variance heterogeneity across batches using a Fligner–Killeen test. The multiplicative batch effect for each IDP was calculated before and after harmonisation. IDPs with *p* > 0.007 were considered to have no significant multiplicative batch effects.

#### 2.5.4 Site-Pairwise batch effects

While the previous omnibus model-comparison test (See 2.5.3 additive batch effect) provides a global assessment of additive batch effects, it may obscure site-specific mean differences between individual batches. To sensitively evaluate batch effects at the level of individual site pairs (total 28 pairs for 8 sites), we fitted a linear mixed-effects model with the (harmonised or raw) IDP as the dependent variable, site as fixed batch effect, biological covariates (age and timepoint), and subject included as a random effect (Eq. 13).

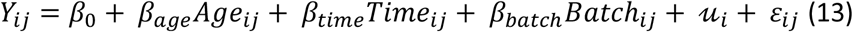

where, 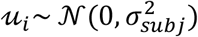 and 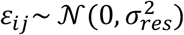.

From this model, we first assessed the overall contribution of site as a fixed effect, testing whether site explained significant variation in mean IDP values. We then conducted pairwise site comparisons using linear contrasts of the fixed-effect site estimates. For each site pair, we tested whether the model-estimated difference in IDP values was significant (p < 0.007, Bonferroni-corrected for seven IDPs).

#### 2.5.5 Multivariate site-wise differences

To quantify multivariate between-site differences, we used the Mahalanobis distance (MD) (Mahalanobis, P. C., 1936), a multivariate extension of Cohen’s *d* (Cohen, 2013) that measures standardized differences between groups (Del Giudice, 2009). MD was included to capture residual site-level divergence in the joint multivariate feature space that may not be detected by univariate batch-effect metrics described above. We first computed a reference distribution using the overall mean and pooled covariance matrix across all batches (Parekh et al., 2022). For each batch, the Mahalanobis distance to the reference was calculated as (Eq. 14):

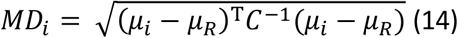

Where *μ*_*i*_ is the mean vector of all IDPs for site *i, μ*_*R*_ is the grand mean vector across sites, and C is the pooled covariance matrix. Larger MD values indicate greater multivariate divergence from the overall distribution. To summarize multivariate site-related heterogeneity, MD values were also averaged across sites. MDs from individual sites and averaged values were compared before and after harmonisation to assess each method’s effectiveness in reducing multivariate between-site differences.

#### 2.5.6 Between-subject variability

To assess whether harmonisation preserves or enhances between-subject variability relative to residual noise, we quantified the proportion of total variance attributable to subject-level differences. Specifically, we fitted a linear mixed-effects model with the IDP (raw and harmonised) as the dependent variable, biological covariates (timepoint and age) as fixed effects, and a subject-specific random intercept to account for repeated measurements (Eq. 12). This model partitions the variance into between-subject variance which reflects inter-individual differences, and residual variance, which captures within-subject variability, measurement noise and unmodelled technical effects. From these variance components, we computed the intraclass correlation coefficient (ICC), defined as the ratio of between-subject variance to total variance (Eq. 15). Under the assumption that harmonisation primarily reduces batch effect without attenuating true biological variability, an increase in ICC after harmonisation indicates an improved relative prominence of genuine between-subject differences.

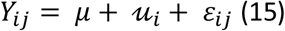

Where, *μ* is fixed intercept, 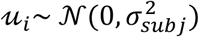 subject-specific random intercept and 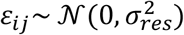 is residual error. From this model, intraclass correlation coefficient (ICC) was calculated as (Eq. 16):

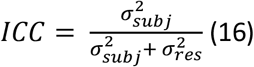

The ICC quantifies the proportion of total variance attributable to stable between-subject differences. Preservation of ICC following harmonisation indicates that subject-level variability is not attenuated, while an increase in ICC suggests improved relative separation of subjects due to a reduction in non-biological residual variance.

#### 2.5.7 Biological variability (age and timepoint)

In this dataset, between-subject biological effects such as age are expected to be preserved, whereas systematic group-level timepoint effects are expected to be small relative to between-subject variability over the short test-retest interval. Evaluating both effects allows assessment of whether harmonisation distorts known biological relationships or introduces spurious longitudinal effects.

To assess preservation of biological variability, we fitted a linear mixed-effects model before and after harmonisation with the IDP (raw and harmonised) as the dependent variable, including age and timepoint as fixed effects and a subject-specific random intercept to account for repeated measurements (Eq. 17):

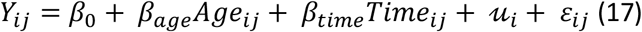

Where, 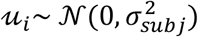 and 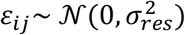.

For each IDP, estimated regression coefficients (*β*), confidence intervals (CIs) (95%) and associated p-values (p-values Bonferroni corrected for 7 IDPs) for age and timepoint were extracted before and after harmonisation. Preservation of age-related associations after harmonisation indicates that biologically plausible between-subject variability is retained, whereas strengthening of age effects may reflect the unmasking of underlying biological trends through removal of site-related confounding or measurement noise. In contrast, marked attenuation or loss of age effects would suggest either removal of meaningful biological signal or reduction of biologically coupled technical variation (e.g., age-related motion). Because no ground-truth biological effect size is identifiable in this pooled dataset that includes inter-scanner acquisitions where different scanners contribute data at different timepoints, changes in the magnitude of age-related effects were interpreted descriptively. In this setting, harmonisation may alter estimated associations by reducing measurement noise or scanner-related confounding, but cannot be assumed to recover a uniquely “true” biological effect. Consequently, our evaluation focused on preservation of biologically plausible associations and consistency across harmonisation methods, rather than absolute effect-size recovery.

At the group level, timepoint effects served as a negative control. Over the short test-retest interval, large and consistent shifts across subjects would be unexpected relative to between-subject differences. The absence of significant timepoint effects before and after harmonisation therefore indicates preserved temporal stability, whereas systematic effects emerging after harmonisation would suggest overcorrection or the introduction of spurious longitudinal variability.

## 3. Results

### 3.1 Variability in raw IDPs

We first examined variability in the raw IDPs across test-retest categories to characterise effects due to scanner heterogeneity prior to harmonisation.

#### 3.1.1 Within-subject variability

**Figure 2a** illustrates the within-subject relative difference between the two timepoints for the normalised brain volume IDP across different test-retest categories: 1) Repeatability (same imaging site and scanner); 2) intra-scanner-model reproducibility (different sites, same scanner model); and 3) inter-scanner reproducibility (different sites and different scanner models). In addition, test-retest differences obtained on the same scanner model are shown separately for GE Signa and Siemens Biograph mMR systems. **Figure 2b** includes a representative slice from the excluded subject (as a result of visual inspection) for the two timepoints, which was removed due to low image quality at one of the timepoints (highlighted in panel a). This example demonstrates that large between-timepoint differences in IDPs can serve as a potential indicator of outlier scans. For other IDPs, see supplementary **Figure S1** for RPD values across subjects.

**Figure 2.**
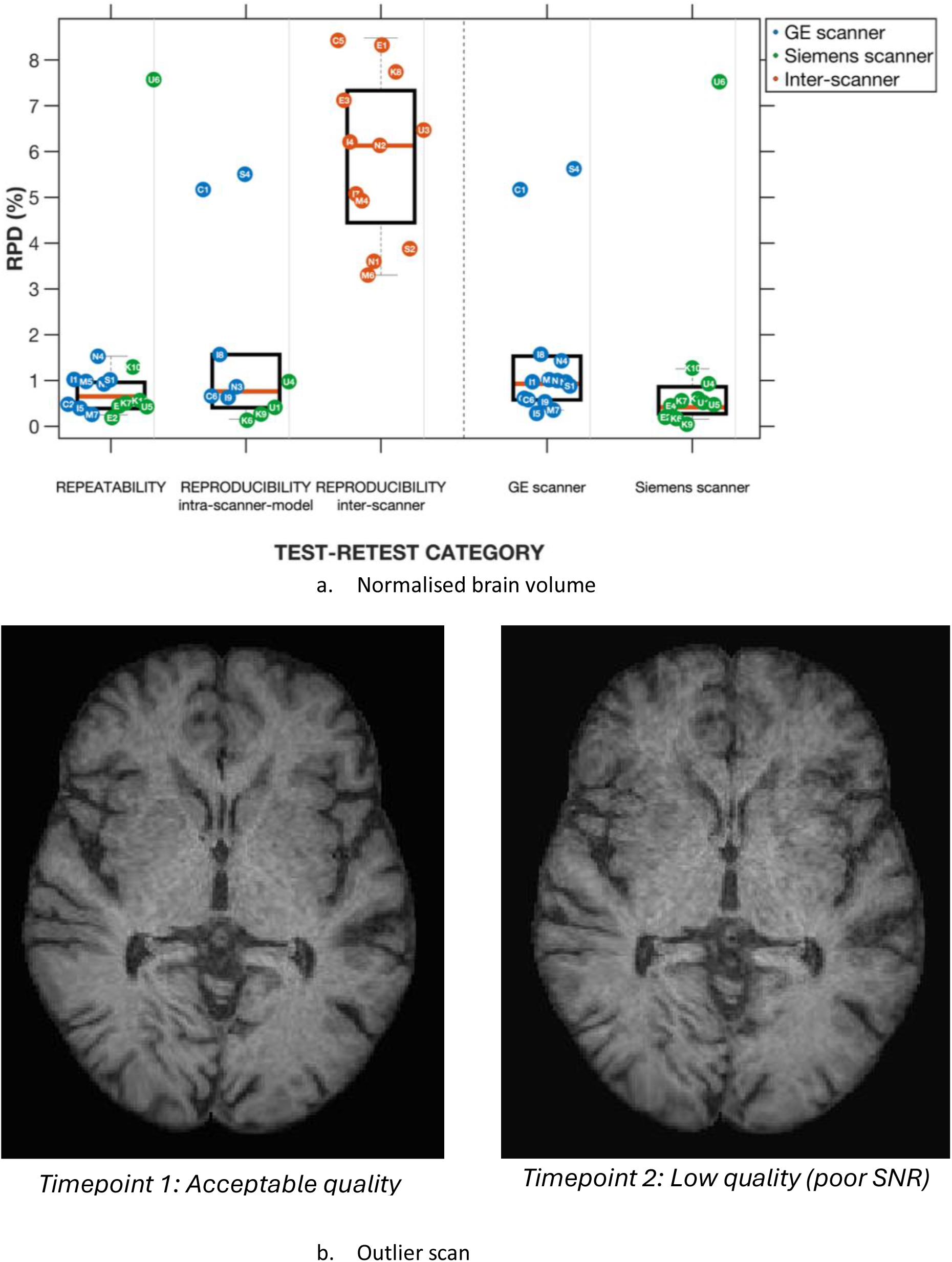
Within-subject IDP differences and visualisation of an outlier scan. Panel a) shows the relative percent difference (RPD, %) in normalized brain volume between two timepoints across test–retest categories: repeatability (same scanner and site), intra–scanner-model reproducibility (same scanner model, different site), and inter-scanner reproducibility (different scanner and site). Intra–scanner-model results (same or different site) are shown separately for GE and Siemens scanners. One participant in the repeatability group exhibited an RPD exceeding that observed in the intra–scanner-model group and comparable to the inter-scanner group; this participant scans are shown in Panel ‘b)’. At timepoint 2, visual inspection revealed poor signal-to-noise ratio and blurring. This participant was excluded from subsequent analyses.

**Figure 3** summarises RPD values across all IDPs for each test-retest category. RPDs were low for repeatability across all IDPs, with median values generally below ∼3% and narrow interquartile ranges (IQRs). Intra-scanner reproducibility showed slightly greater dispersion than repeatability, though median RPDs remained modest across tissues. In contrast, inter-scanner reproducibility demonstrated substantially higher variability, with markedly elevated median RPDs and wider IQRs, particularly in WM and CSF. WM in this group exhibited the largest spread, with several outliers exceeding 20–30%, indicating pronounced between-scanner variability. Scanner-wise analyses (GE and Siemens) demonstrated consistently low median RPDs with overlapping distributions and limited dispersion. Overall, variability followed a clear gradient of increasing RPD from repeatability to intra-scanner reproducibility to inter-scanner reproducibility, with tissue-specific effects most pronounced in WM and CSF.

**Figure 3.**
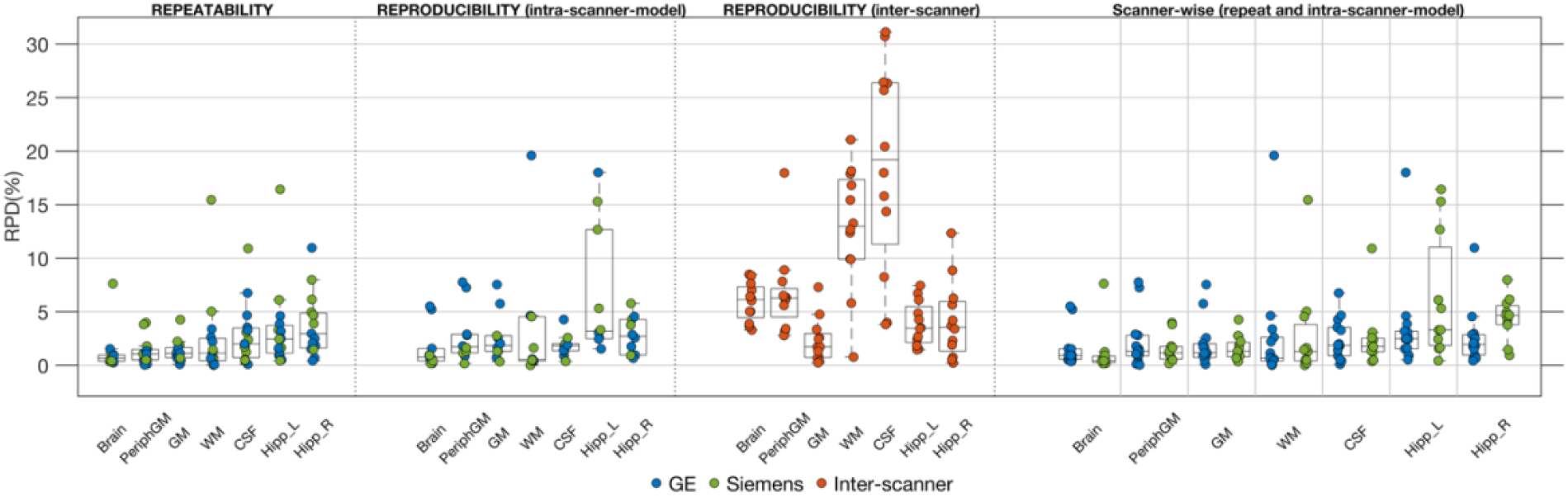
Within-subject relative percent difference for each IDP between the two timepoints across the three test-retest categories: repeatability (same scanner and site), intra–scanner-model reproducibility (same scanner model, different site), and inter-scanner reproducibility (different scanner and site). Intra– scanner-model results (same or different site) are shown separately for GE and Siemens scanners.

#### 3.2.2 Subject order consistency

As shown in **Figure 4**, the repeat-scanner group exhibited the strongest preservation of subject rank ordering between timepoints, with Spearman correlations exceeding 0.9 across all IDPs. Under the permutation test of random subject ordering, all repeat-scanner correlations were statistically significant (*p* < 0.05), indicating rank preservation that was unlikely to arise by chance.

**Figure 4.**
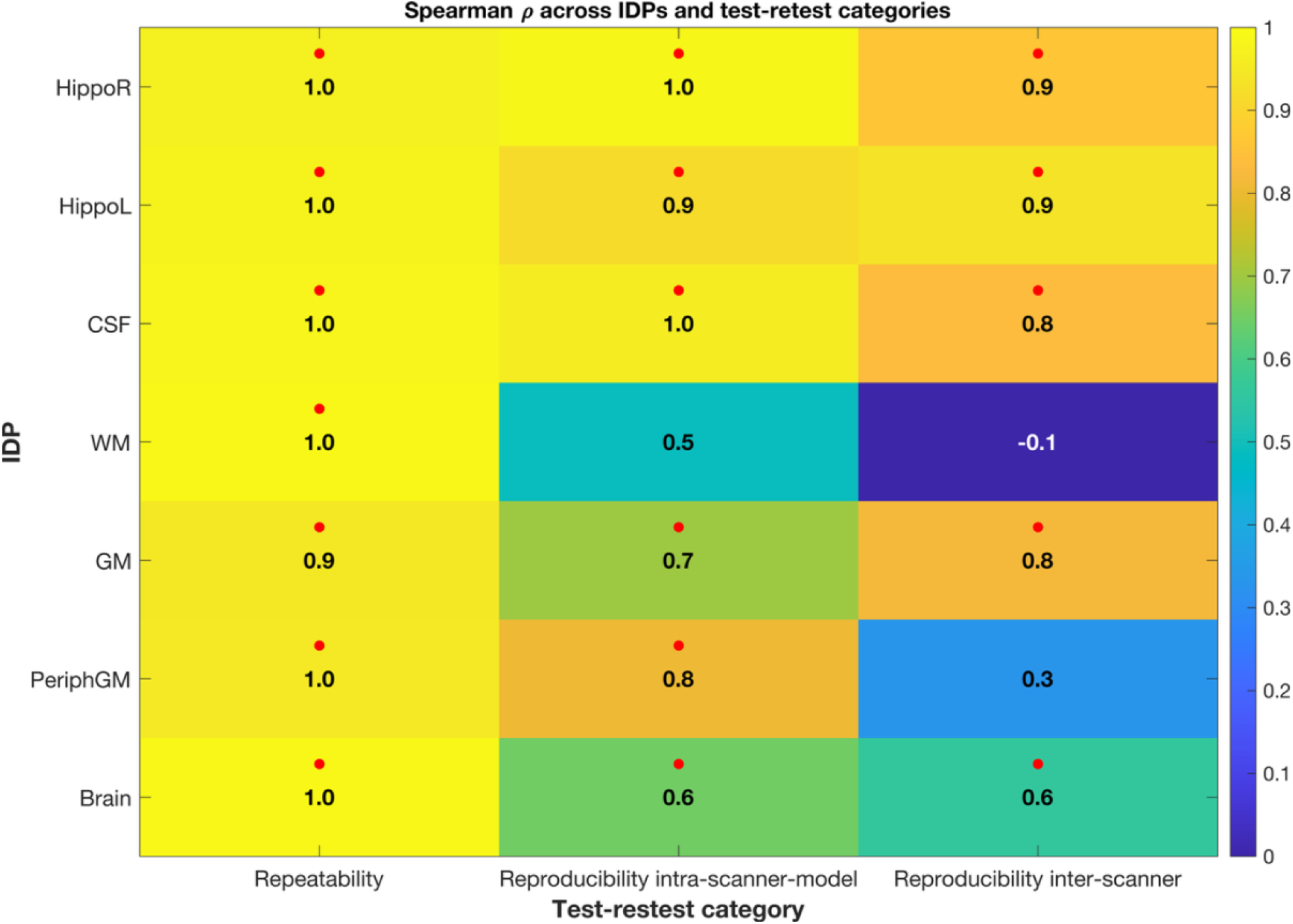
Spearman rank correlation coefficients (ρ) between timepoints for each IDP and test-retest category (repeatability (same scanner and site), intra–scanner-model reproducibility (same scanner model, different site), and inter-scanner reproducibility (different scanner and site)). Heatmap cells show observed ρ (higher = better preservation of subject ranking). The p-value from permutation test equals the proportion of permuted correlations with absolute value ≥ the observed correlation. A significant result (p<0.05) highlighted with red coloured marker indicates that the observed correlation is greater than expected under random subject ordering.

In the intra-scanner group, subject order consistency varied across IDPs. Spearman rank correlation coefficient was the lowest for WM (ρ = 0.5, *p* > 0.05), and moderate for Brain (ρ = 0.6, *p* < 0.05) and GM (ρ = 0.7, *p* < 0.05), whereas CSF and hippocampal volumes demonstrated high consistency (ρ ≈ 0.9–1.0, *p* < 0.05).

Inter-scanner subject order consistency varied substantially across IDPs. Total brain (ρ = 0.6, *p* < 0.05) and GM (ρ = 0.8, *p* < 0.05) IDPs demonstrated moderate-to-high agreement, whereas peripheral GM showed low consistency (ρ = 0.3, *p* > 0.05). WM exhibited poor agreement (ρ = −0.1, *p* > 0.05), indicating substantial rank instability across scanners. In contrast, CSF and hippocampal volumes showed relatively high baseline consistency (ρ ≈ 0.8–0.9, *p* < 0.05).

Taken together, in the raw data, both within-subject RPDs and subject rank consistency differed systematically across test-retest categories, with greater variability and reduced timepoint-wise correspondence observed under increasing scanner heterogeneity. In the following sections, we report the effects of applying harmonisation methods on these measures and present additional evaluation metrics that explicitly model batch structure in pooled dataset.

### 3.2 Harmonisation evaluation

We visually observed the overall distribution of IDPs before and after harmonisation for the pooled dataset. **Figure 5** illustrates the distributions of normalised total brain volume across batches before and after harmonisation (Refer to supplementary section (**Figure S2**) for similar plots for other IDPs). Prior to harmonisation (panel a) ‘Raw’), the batches with Siemens sites (EDI, KCL, UCL) consistently showed higher brain volumes compared to the batches with GE sites (CAM, ICL, MAN, NEW, SHF), reflecting clear scanner-related differences. After harmonisation, the scanner distributions were shifted toward the global mean across subjects (dash-dot (--) line in each subplot), indicating that each method reduces these systematic batch effects to varying degrees. These plots provide an intuitive overview of the impact of harmonisation on batch-related differences, illustrating that different methods reduce systematic batch effects to varying extents. In the next section, we quantify these effects in more detail using the proposed evaluation metrics.

**Figure 5.**
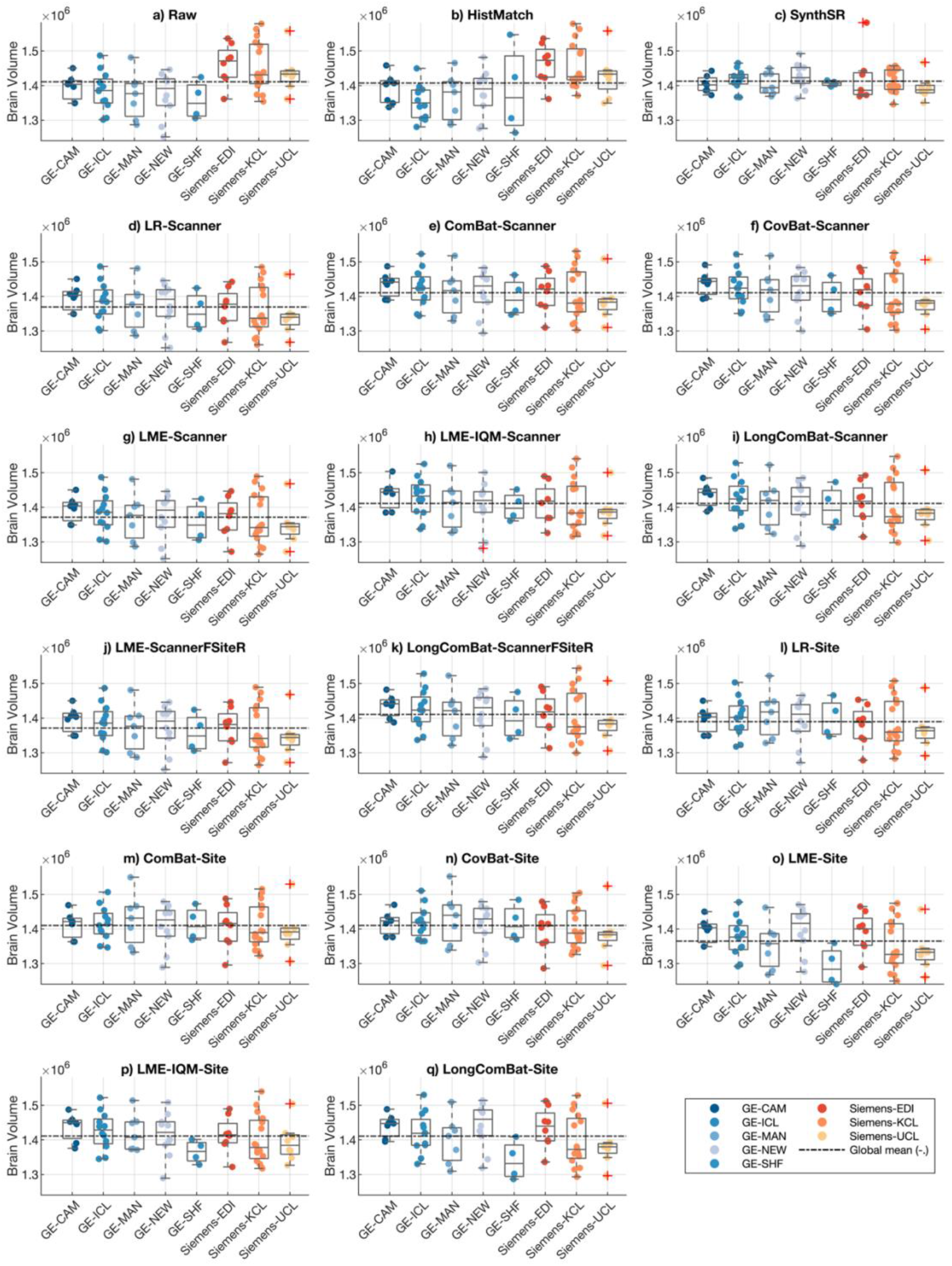
Distribution of normalised total brain volume across different sites and scanners before and after applying image and statistical harmonisation methods to the dataset

#### 3.2.1 Within-subject variability

Harmonisation effects were evaluated by recalculating relative percent differences after application of different harmonisation methods and statistically comparing medians of the difference against the unharmonised (‘Raw’) values using non-parametric Wilcoxon rank-sum tests (**Figure 6**). Median RPD values from repeatability group (‘RepeatRaw’) are shown as a reference benchmark to assess whether harmonisation reduced reproducibility error in intra and inter-scanner categories toward the repeatability floor. For more details on the RPD values from each participant, IDP, group and method, see supplementary materials **Figure S1** and excel sheet – ‘*within_subject_variability*’; also see the previous section (3.1.1) for RPD values before harmonisation.

**Figure 6.**
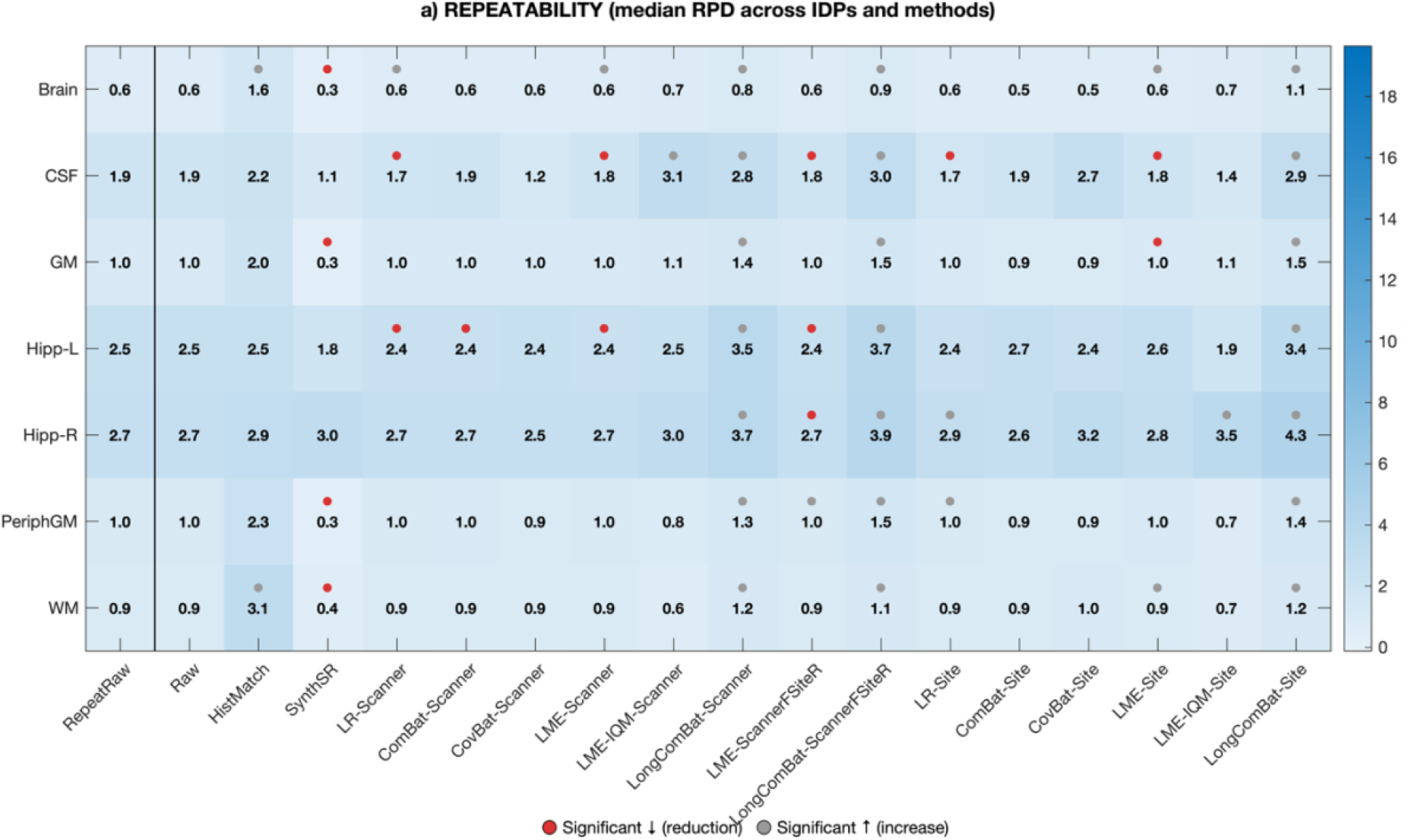
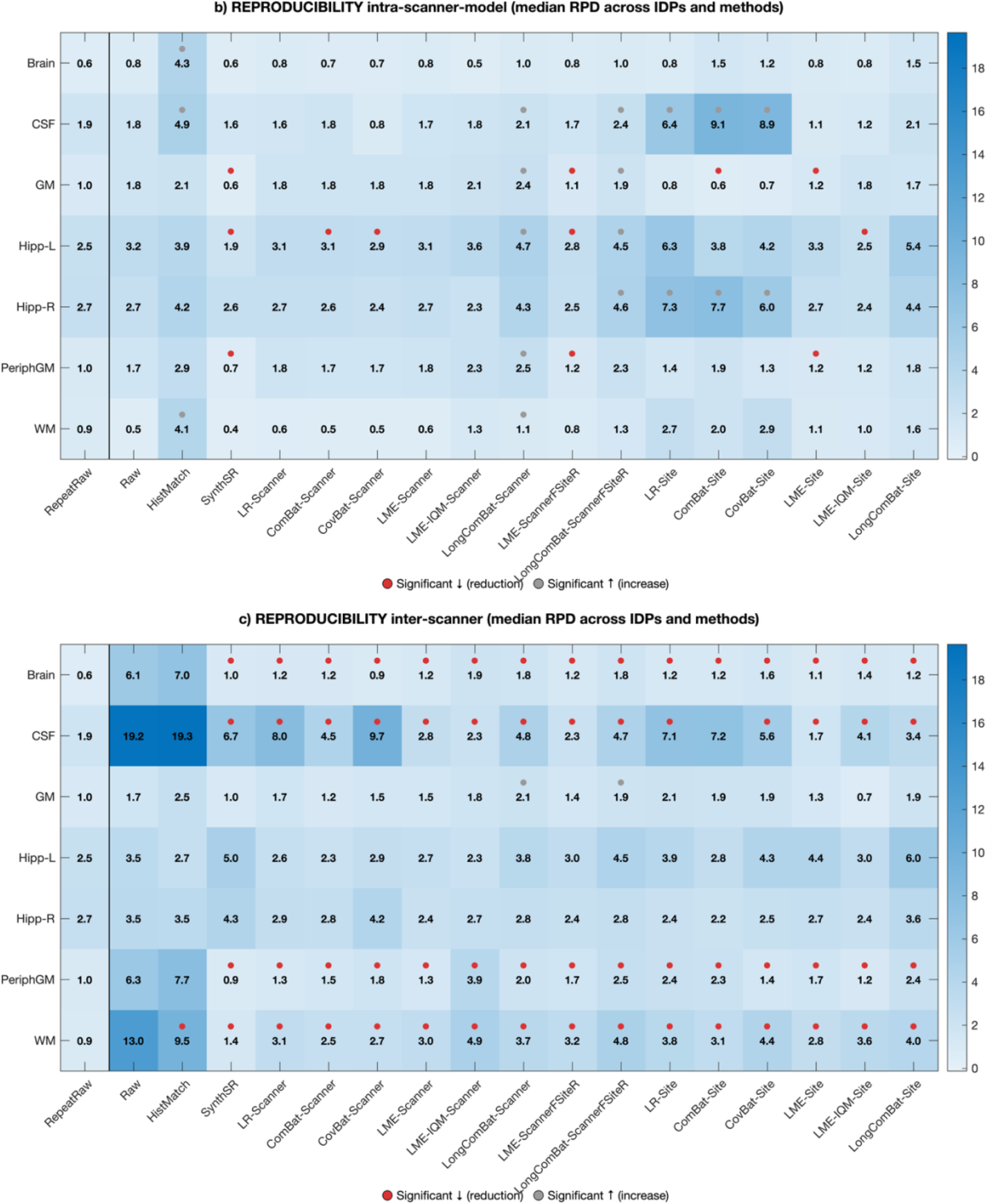
Median values of relative percent difference (RPD) before (‘Raw’ on x-axis) and after applying harmonisation methods. The heatmaps display the median relative percent difference (RPD) between timepoints for each imaging-derived phenotype (IDP; rows) and harmonisation method (columns) across three test–retest settings: (a) Repeatability, (b) Reproducibility (intra-scanner model), and (c) Reproducibility (inter-scanner). The leftmost column (‘RepeatRaw’ on x-axis) represents the baseline median RPD in the repeatability group and serves as an estimate of the technical noise floor. This baseline enables comparison of whether harmonisation in the intra- and inter-scanner settings reduces RPD toward repeatability levels. Colored markers indicate statistically significant paired differences relative to the raw data within each group (Wilcoxon signed-rank test, α = 0.05). Red markers denote a significant reduction in RPD (improved measurement stability), whereas grey markers denote a significant increase in RPD (deterioration in within-subject consistency).

##### Repeatability group

As shown in **Figure 6a**, the image-based harmonisation approach using histogram matching (HistMatch) showed significant increase in RPD in some IDPs (total brain, WM), whereas the SynthSR method had minimal impact and preserved low variability in most IDPs (ranging from 0.3-3%). For statistical harmonisation methods, those using either scanner or mixed batch formulation similarly maintained RPDs at baseline levels (except variants of Longitudinal ComBat). Among methods using site as batch effect, both cross-sectional (LR-Site, ComBat-Site, CovBat-Site) and those incorporating repeated measures structure (LME-Site, LME-IQM-Site) showed negligible effects (except Longitudinal ComBat increased RPD values significantly). Overall, no harmonisation strategy meaningfully improved repeatability, indicating that the repeatability floor reflects intrinsic measurement stability rather than correctable scanner bias.

##### Intra-scanner reproducibility

As shown in **Figure 6b**, raw intra-scanner RPDs were already close to repeatability levels (ranging from 0.5-3.2%). The image-based harmonisation approach using histogram matching increased variability across IDPs (significant increase in total brain, CSF and WM IDPs from Raw), while SynthSR significantly reduced RPD values in some IDPs even below the repeatability floor (GM, Peripheral GM and left hippocampus IDPs). For statistical harmonisation methods, those using either scanner or mixed batch formulation largely preserved baseline RPDs, with only modest tissue-specific changes except longitudinal ComBat variants which significantly increased RPD values relative to Raw. A distinction emerged within the methods using site as batch effect: cross-sectional approaches (LR-Site, ComBat-Site, CovBat-Site) showed variable performance and inflation of median RPD values (e.g., CSF, right hippocampus IDPs), whereas other methods incorporating repeated structure (LME-Site, LME-IQM-Site, LongComBat-Site) comparatively demonstrated greater stability and avoided this inflation. However, because baseline intra-scanner variability was already low, harmonisation provided limited additional benefit in this group.

##### Inter-scanner reproducibility

As shown in **Figure 6c**, raw RPD values were substantially elevated, particularly in CSF (median – 19.2%) and WM IDPs (median – 13%). Image-based harmonisation approach using histogram matching (HistMatch) yielded partial and tissue-dependent reductions but did not consistently approach repeatability levels. In contrast, SynthSR achieved stronger improvements in some IDPs (total brain, peripheral GM and CSF), though performance remained variable for, e.g., median RPD values increased for hippocampus IDPs (not statistically significant). Statistical harmonisation methods using either scanner or mixed batch formulation produced the most consistent and substantial reductions across tissues, markedly lowering CSF and WM RPDs and bringing whole-brain and GM measures close to the repeatability floor. Within the methods using site as batch effect, cross-sectional methods (LR-Site, ComBat-Site, CovBat-Site) reduced RPD relative to Raw, e.g., significant reduction in total brain, peripheral GM and WM IDPs. Methods using site as batch effect (LME-Site, LME-IQM-Site, LongComBat variants) achieved significant reductions across several IDPs, though their performance remained IDP-dependent. These findings indicate that inter-scanner variability is largely systematic and most effectively mitigated when harmonisation explicitly models scanner effects or incorporates longitudinal structure.

Comparison of variability from intra- and inter-scanner groups against the repeatability benchmark (‘RepeatRaw’) demonstrated that no harmonisation method uniformly restored inter-scanner RPDs to the repeatability floor across all IDPs. Whole-brain and GM IDPs most consistently approached repeatability levels under several statistical methods, whereas CSF and hippocampal volumes remained elevated despite substantial reductions. WM showed marked improvement, particularly with SynthSR and statistical approaches, though complete convergence to repeatability was not consistent across methods.

#### 3.2.2 Subject order consistency

Spearman rank correlations indicating subject order consistency for each IDP across two timepoints for 3 groups before and after applying each harmonisation method are summarised in **Figure 7**. For more details on the correlation values from each IDP, group and method, see supplementary excel sheet – ‘*subject_order_consistency*’; also see the previous section (3.1.2) for correlation values on data before harmonisation.

**Figure 7.**
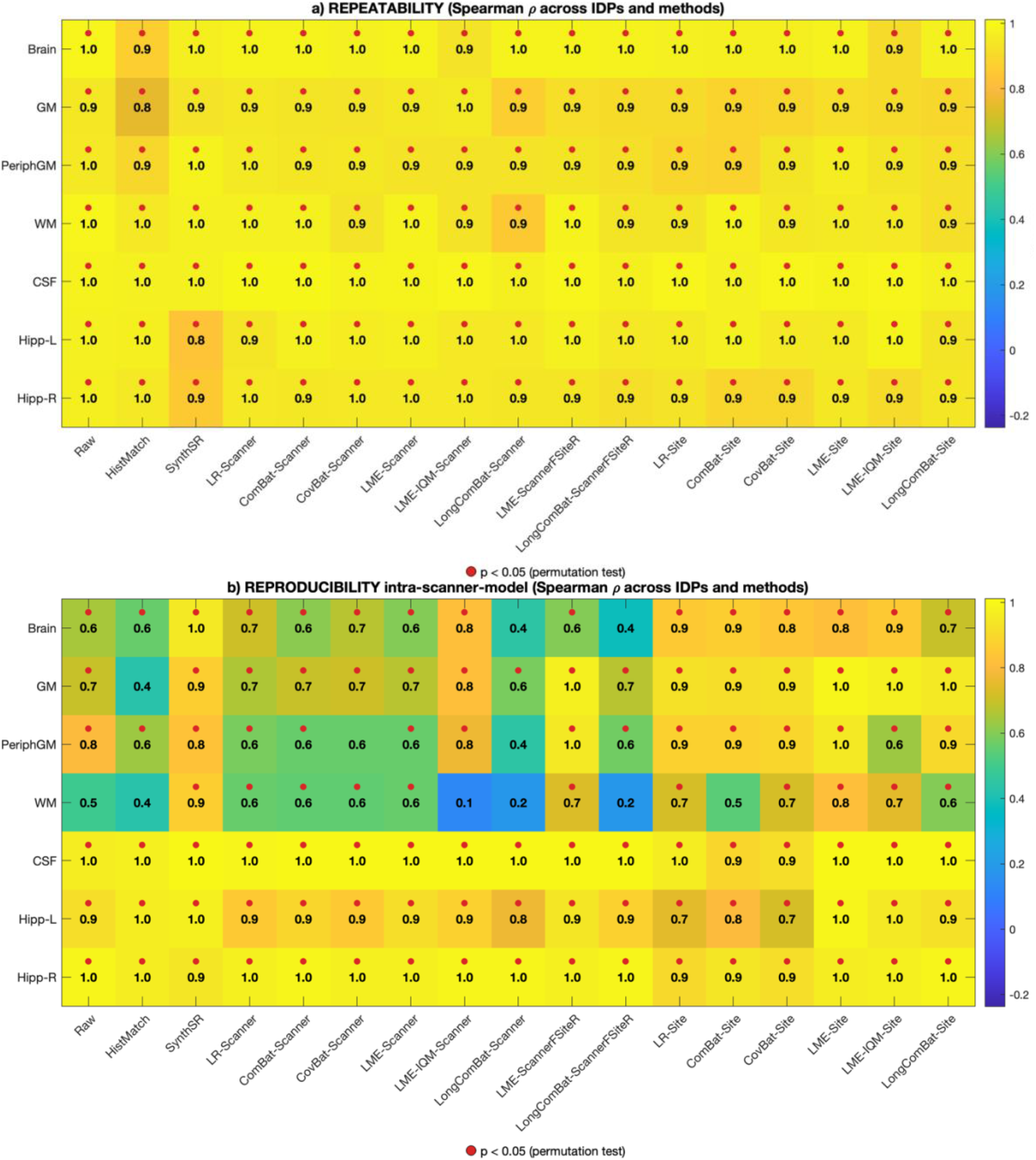
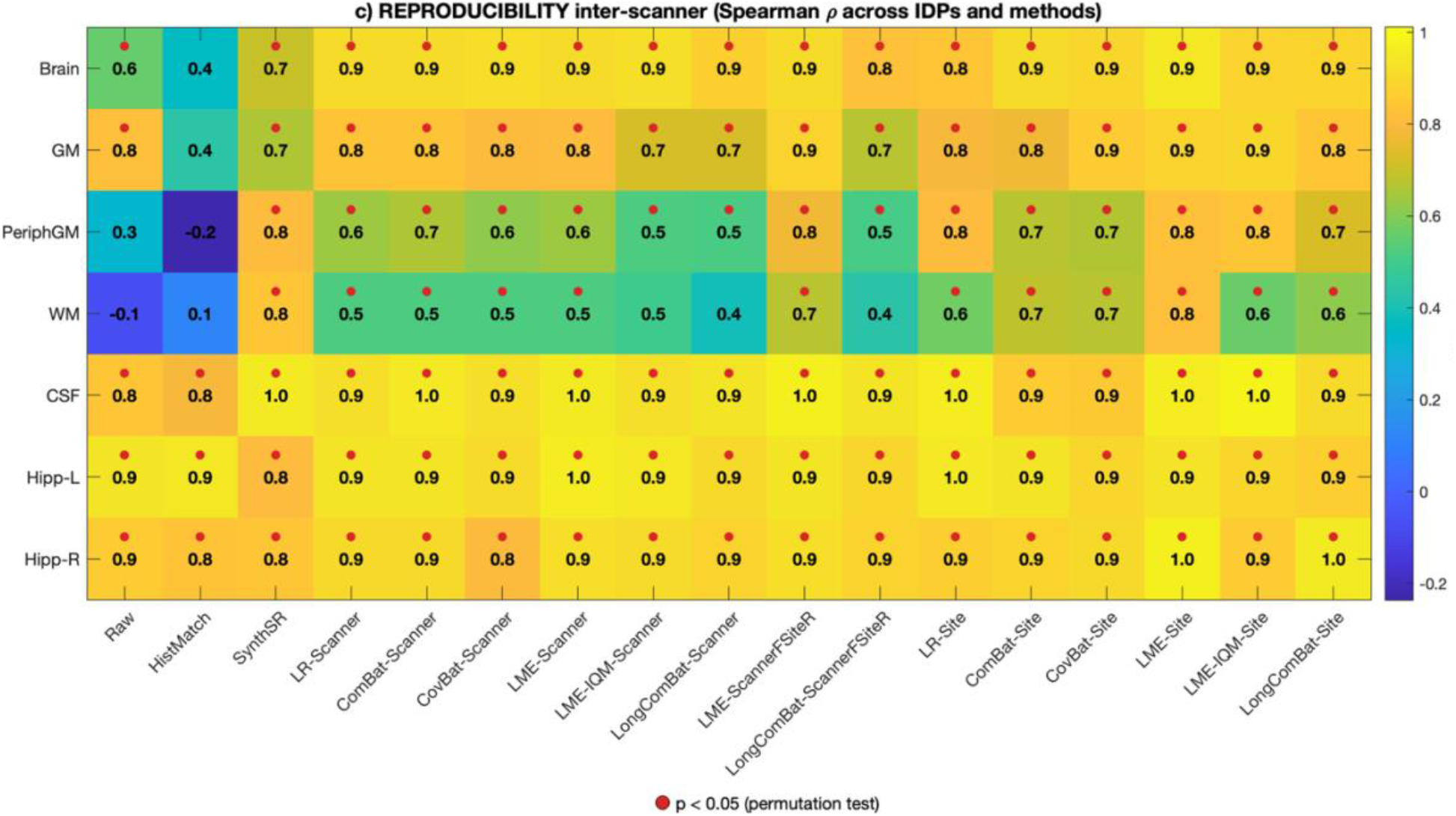
Subject-order consistency before (‘Raw’ on x-axis) and after applying harmonisation methods: Heatmaps showing Spearman rank correlation coefficients assessing subject-order consistency for each IDP (rows) and harmonisation method (columns) across three test-retest categories: a) Repeatability, b) Reproducibility intra-scanner model, and c) Reproducibility inter-scanner groups. Cell values represent Spearman ρ, red markers indicate statistically significant correlations (permutation test, *p* < 0.05). Values closer to 1 indicate stronger preservation of inter-subject ranking consistency, whereas values near 0 indicate lower/reduced rank consistency across timepoints.

##### Repeatability group

As shown in **Figure 7a**, Overall excellent subject order consistency was observed even before harmonisation (ρ ≈ 0.9-1.0) in the repeatability group with only very slight reductions for some IDPs after applying harmonisation methods.

##### Intra-scanner reproducibility

As shown in **Figure 7b**, following image-based harmonisation using histogram matching, subject order consistency was further reduced for GM and WM (both ρ ≈ 0.4), indicating decreased rank stability relative to the unharmonised data. In contrast, SynthSR-based harmonisation increased ordering consistency across all IDPs (ρ ≥ 0.8), with notable improvements in Brain and WM measures compared to unharmonised (‘Raw’) data. Statistical harmonisation approaches modelling scanner as the batch effect generally reduced intra-scanner consistency, particularly for peripheral GM (ρ ≈ 0.6). Methods incorporating repeated measures structure (e.g., LME-IQM-Scanner, LongComBat-Scanner, LongComBat-ScannerFSiteR) resulted in marked reductions in WM ordering (ρ ≈ 0.1–0.2), indicating substantial rank disruption. Conversely, methods modelling site as the batch effect generally preserved or improved intra-scanner ordering across IDPs. These approaches achieved high consistency (ρ ≥ 0.9), particularly for global GM and hippocampal volumes, suggesting greater stability of subject-level ranking.

##### Inter-scanner group

As shown in **Figure 7c**, Histogram matching did not improve inter-scanner subject rank ordering and further reduced consistency for several IDPs (e.g., Brain ρ = 0.4; peripheral GM ρ = −0.2; WM ρ = 0.1), indicating persistent rank distortion. SynthSR harmonisation markedly improved ordering consistency across all IDPs, with correlations ≥ 0.7 across all IDPs, restoring strong monotonic agreement between scanners. Statistical harmonisation approaches modelling scanner as the batch effect generally improved inter-scanner consistency relative to Raw, particularly for Brain (ρ ≈ 0.9) and peripheral GM (ρ ≈ 0.6–0.7). However, improvements for WM were moderate (ρ ≈ 0.5), and other methods (e.g., LME-IQM-Scanner, LongComBat-Scanner variants) yielded only partial recovery (ρ ≈ 0.4–0.5). Methods modelling site as the batch effect consistently produced strong inter-scanner agreement across IDPs. These approaches achieved high correlations for Brain and GM (ρ ≈ 0.8–0.9), improved WM consistency (ρ ≈ 0.6–0.8), and maintained high agreement for CSF and hippocampus volumes (ρ ≈ 0.9–1.0), suggesting robust correction of scanner-induced rank distortion under these methods.

Building on the subject-level analyses in Sections 3.2.1 and 3.2.2, we next evaluated residual batch effects at the site level using a common site-based evaluation framework across all harmonisation methods. Although the methods modelled batch effects using different formulations (scanner, site, or mixed), all outcomes were assessed with respect to site to provide a consistent and conservative comparison.

#### 3.2.3 Additive batch effect

As shown in **Figure 8**, significant additive batch effects attributable to site were observed prior to harmonisation for all IDPs (Raw), with the exception of left hippocampal volume, which did not survive multiple-comparison correction. Following image-based harmonisation, additive batch effect largely persisted for histogram matching (HistMatch) which failed to remove additive effect for most IDPs, whereas SynthSR successfully eliminated additive batch effects for the majority of IDPs, with residual effects observed for CSF and right hippocampus volumes. After applying harmonisation methods under different batch-modelling formulations, when scanner was modelled as the batch effect, none of the methods successfully removed additive site effect across all IDPs. In methods using mixed batch formulations (treating scanner as a fixed effect and site as a random effect), only LME method (LME-ScannerFSiteR) significantly removed site effects from all IDPs. When site was explicitly modelled as the batch effect, LME (LME-Site), IQM method (LME-IQM-Site), and longitudinal ComBat (LongComBat-Site) successfully eliminated additive batch effect from all IDPs. The p-values for all IDPs, methods are provided in supplementary excel sheet – *‘additive_batch_effect*’).

**Figure 8.**
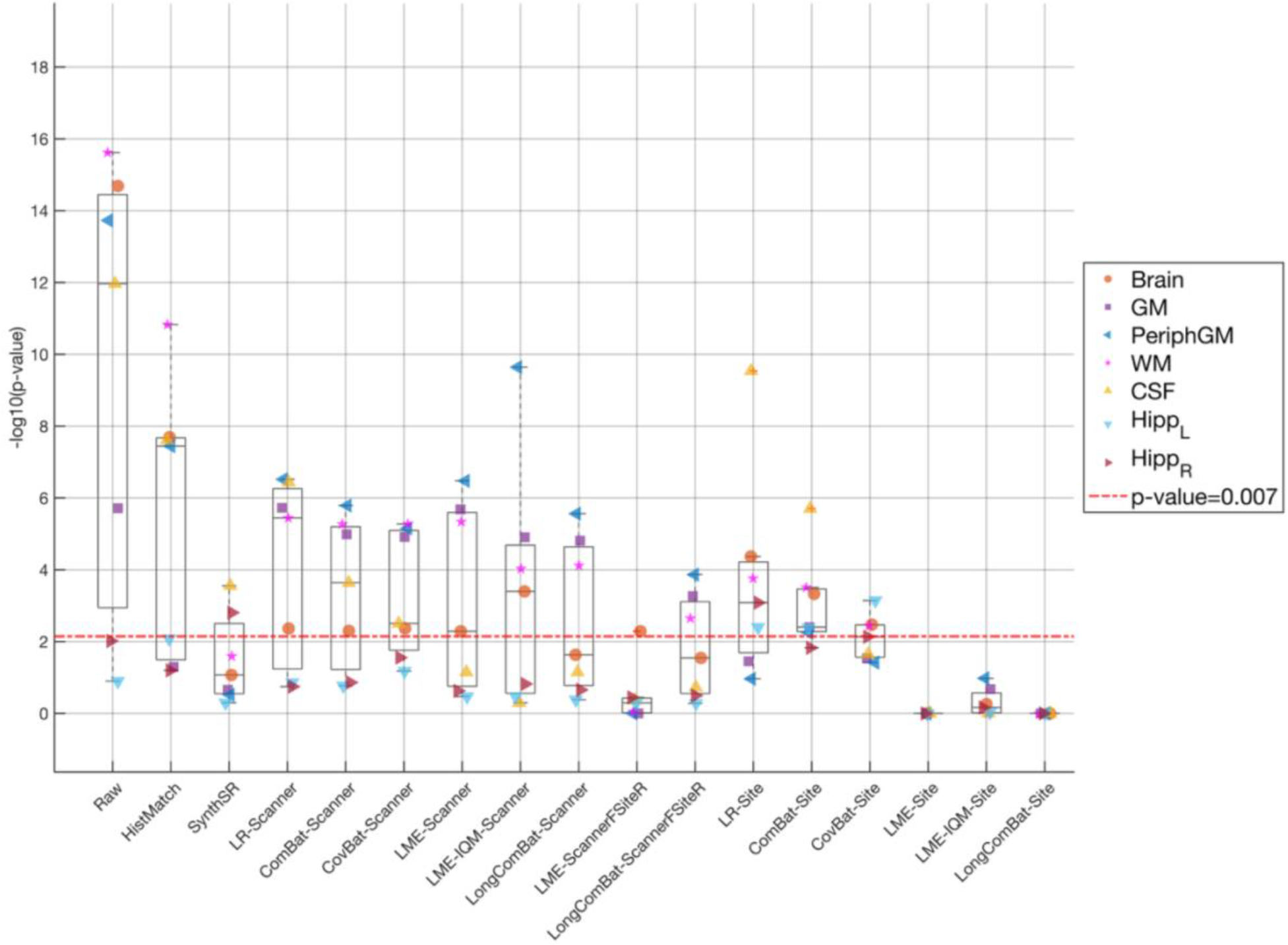
Significance tests for additive (mean-shift) site batch effect for each IDP before and after harmonisation. The *p*-values were obtained from Kenward–Roger F-tests comparing linear mixed-effects models with and without site as a fixed effect. Values are shown as −log10(*p*), with the dotted red line indicating the Bonferroni-corrected significance threshold for seven IDPs (*p* = 0.007). IDPs above the threshold denote statistically significant additive batch effect.

#### 3.2.4 Multiplicative batch effect

In contrast to additive effects, multiplicative (scaling) batch effects were absent prior to harmonisation across most IDPs (**Figure 9**). Following harmonisation, multiplicative batch effects remained non-significant across IDPs for all the harmonisation methods. The p-values for all IDPs, methods are provided in supplementary excel sheet – *‘multiplicative_batch_effect*’).

**Figure 9.**
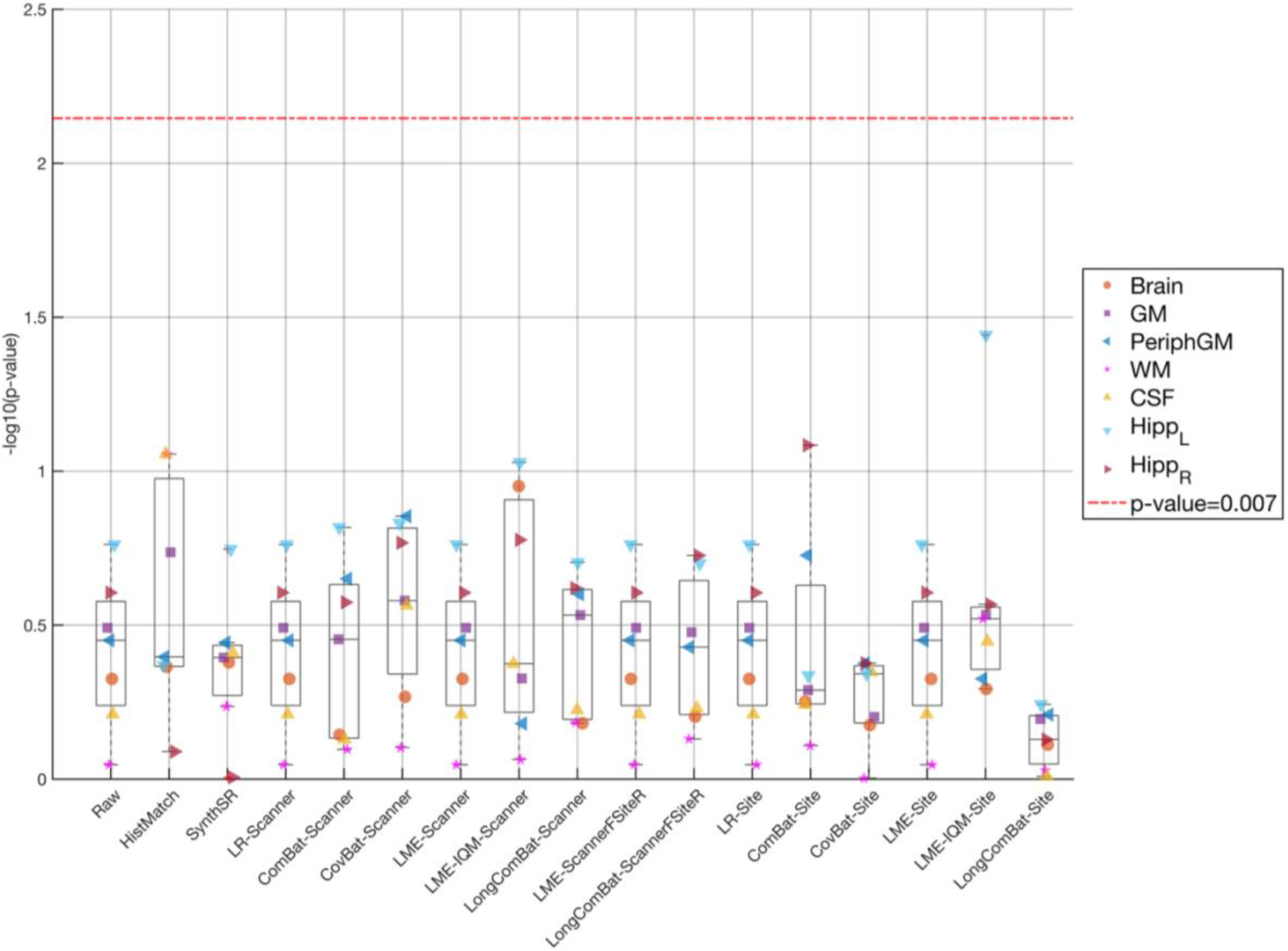
Significance tests for multiplicative (scaling) site batch effect for each IDP before and after harmonisation after excluding the site with 2 samples. The *p*-values were obtained from Fligner–Killeen tests applied to model residuals across sites. Values are shown as −log10(*p*), with the dotted red line indicating the Bonferroni-corrected significance threshold for seven IDPs (*p* = 0.007). IDPs above the threshold denote statistically significant multiplicative batch effects.

#### 3.3.5 Site-pairwise batch effect

We next examined site-specific mean differences using fixed-effect site models and pairwise contrasts, which assess differences in mean IDP values between sites rather than the contribution of site to overall variance assessed in the previous additive batch effect analysis. As shown in **Figure 10** (also see supplementary excel sheet – ‘*site_pairwise_batch_effect*’), before harmonisation (Raw), the *overall* fixed effect of site was significant *across* most of the IDPs (except left hippocampus), indicating substantial site-related batch effect. Pairwise site contrasts revealed widespread site-specific differences, with the number of significant site pairs (out of 28 possible comparisons) ranging from 1 for left hippocampal volume to 20 for whole-brain and peripheral GM volumes.

**Figure 10.**
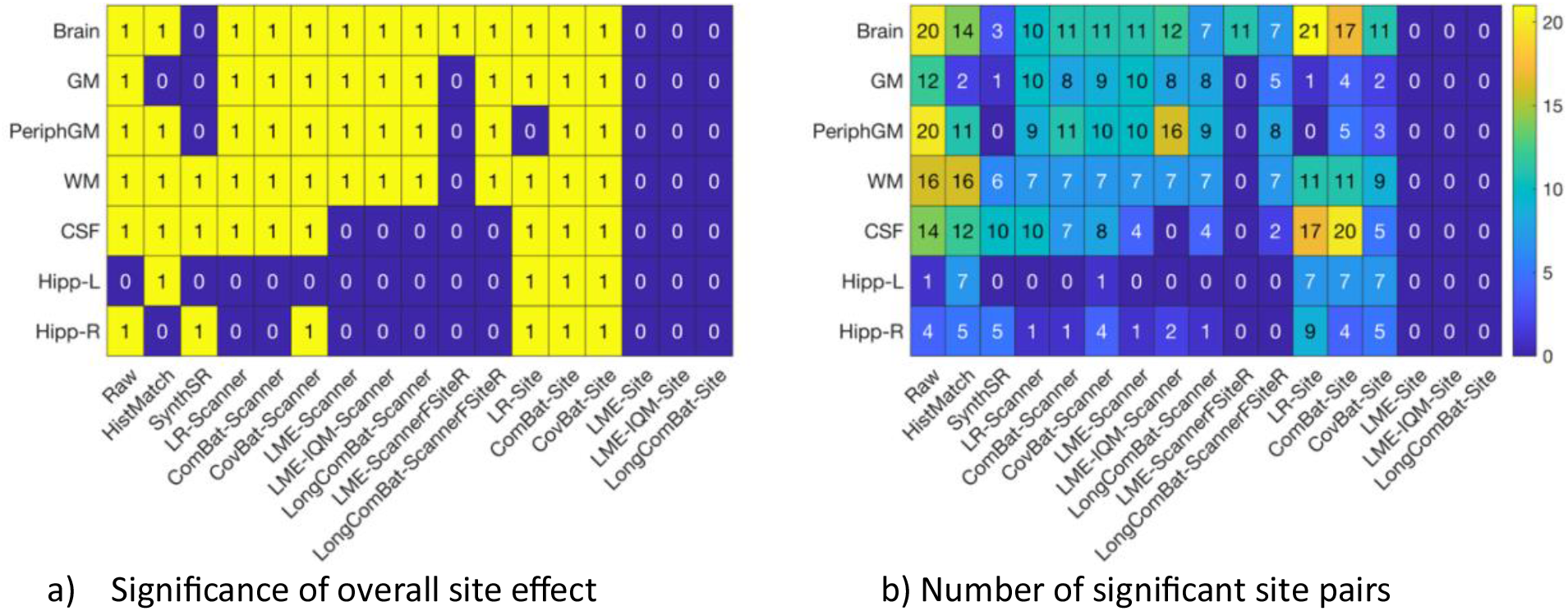
Site-pairwise batch effects before and after harmonisation across all IDPs. a) Significance of the overall fixed effect of site from linear mixed-effects evaluation model (1: *p* < 0.007; 0: *p* ≥ 0.007), b) Number of site pairs exhibiting significant differences for each IDP based on linear contrasts of fixed-effect site estimates (total site pairs = 28).

Following image-based harmonisation, the overall fixed effect of site was significant for most IDPs after histogram matching (HistMatch), although the number of significant site pairs was reduced for some IDPs in comparison to raw data, except for WM and hippocampus volumes. In contrast, SynthSR substantially reduced site-pairwise batch effects. The overall fixed effect of site was no longer significant for total brain, GM, peripheral GM, and left hippocampus volumes, and the number of significant site pairs was markedly reduced across IDPs. After applying harmonisation methods under different batch-modelling formulations, approaches using scanner as the batch variable generally failed to eliminate the overall fixed effect of site for most IDPs but consistently reduced the number of significant site-pair differences. Among mixed batch formulations, only LME (LME-ScannerFSiteR) successfully removed the overall site effect for most IDPs and eliminated all significant site-pair differences (except total brain volume). Methods explicitly modelling site as the batch effect (LME-Site, LME-IQM-Site, and LongComBat-Site) eliminated statistically significant global site effects, with no site-pair comparisons remaining significant across any IDP.

#### 3.3.6 Multivariate site-wise differences

Mahalanobis distances were used to quantify multivariate deviations of each site from the pooled reference distribution defined by all sites combined (**Figure 11**) (also see supplementary excel sheet – ‘*multivariate_variability*’). Before harmonisation (Raw), Mahalanobis distances ranged from 1.4 to 3.2 across sites, with an average distance of 2.4. The largest deviation from the reference distribution was observed for the ICL–GE site (3.2), whereas the smallest deviation was observed for the KCL–Siemens site (1.4), indicating substantial site-specific multivariate differences in the before harmonisation.

**Figure 11.**
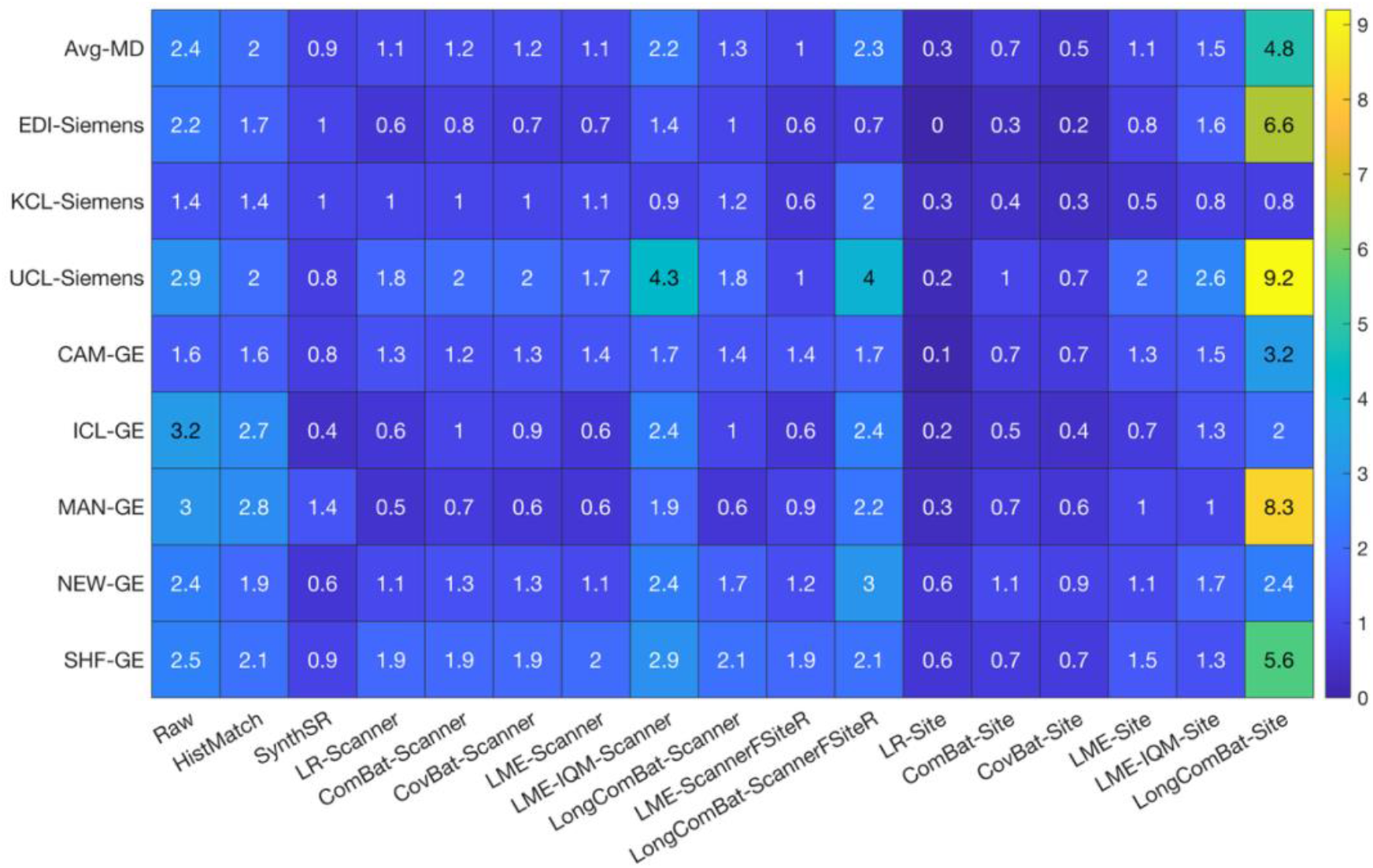
Mahalanobis distances (MDs) across sites before and after harmonisation. Each value for each site and method indicate the multivariate distance of that site from the reference distribution except the *first* row which shows summarised average MD across all sites for each method.

After applying image-based harmonisation methods, histogram matching (HistMatch) produced only modest reductions in Mahalanobis distance across sites (average 2.0), indicating limited attenuation of multivariate site differences. In contrast, SynthSR substantially reduced Mahalanobis distances across all sites, with an average distance of 0.9, indicating strong alignment of sites with the pooled reference distribution. Statistical harmonisation methods using scanner as the batch variable reduced Mahalanobis distances for most sites; however, distances increased for the UCL–Siemens site. LME method (LME-ScannerRSiteR) with mixed batch formulations (scanner as fixed effect and site as random effect) performed more robustly and consistently reduced Mahalanobis distances across all sites, achieving an average distance of 1.0. When site was explicitly modelled as the batch effect, most methods reduced Mahalanobis distances across sites; however, longitudinal ComBat increased distances for most sites, resulting in the highest average distance among all harmonisation approaches (4.8).

#### 3.3.7 Between-subject variability

**Figure 12** summarises between-subject variability quantified using intraclass correlation coefficients (ICC) before and after harmonisation (see supplementary excel sheet ‘*between_subject_variability*’ for ICC values across all IDPs and methods). Prior to harmonisation (Raw), ICC values were high (>0.9) for most IDPs, indicating that a large proportion of variance reflected between-subject differences. Lower ICC values were observed for total brain (0.7), peripheral GM (0.6), and WM (0.5) volumes, indicating a reduced proportion of between-subject variability relative to within-subject variability for these IDPs.

**Figure 12.**
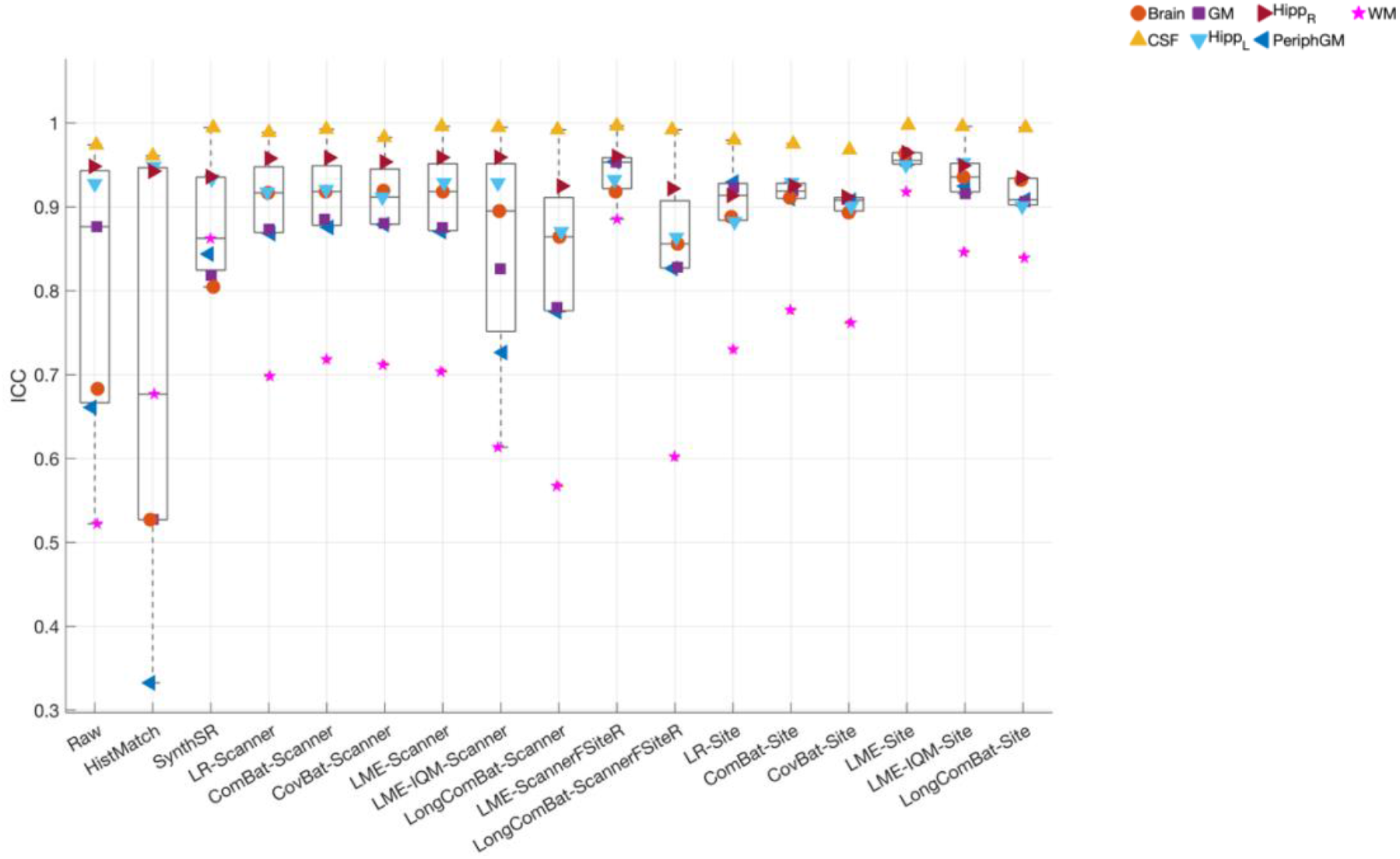
Intraclass correlation coefficients (ICCs) before and after harmonisation for IDPs.

After applying image-based harmonisation methods, histogram matching did not improve between-subject variability and, in several cases, further reduced ICC values, particularly for total brain (0.5), GM (0.5), and peripheral GM (0.3) volumes. In contrast, SynthSR increased ICC for most IDPs, indicating improved preservation of between-subject variability following harmonisation, although a modest reduction was observed for GM volumes (0.8).

After applying statistical harmonisation, methods using scanner as the batch variable improved ICC for most IDPs. LME method (LME-ScannerFSiteR) with mixed batch formulations showed more consistent and increased ICC across all IDPs, although longitudinal ComBat (LongComBat-ScannerFSiteR) provided only modest improvement for WM volumes. Methods explicitly modelling site as the batch effect produced consistent improvements in ICC across all IDPs.

#### 3.3.8 Biological variability

##### Association between IDPs and Age

**Figure 13** shows associations between age and IDPs estimated using linear mixed-effects models before and after harmonisation (see supplementary excel sheet ‘*biological_variability_Age_IDP*’ for more details). Age-IDP associations were directionally consistent across harmonisation strategies. For GM IDP, negative associations were observed in the Raw data (β = −0.4) and remained stable following statistical harmonisation (typically β = −0.4 to −0.5, frequently Bonferroni-significant). Hippocampus volumes showed strong negative associations with age in Raw data (Hipp-L β = −0.6; Hipp-R β = −0.7) that were preserved across all methods (β ≈ −0.5 to −0.7, *p* < 0.007). CSF demonstrated positive associations that increased following harmonisation (Raw β = 0.2; harmonised β ≈ 0.3–0.4), with significance achieved under most statistical approaches. Whole-brain and peripheral GM effects were modest in Raw data (Brain β ≈ 0.0; PeriphGM β = −0.2) but became more consistently negative after harmonisation (Brain β ≈ −0.4; PeriphGM β ≈ −0.4 to −0.5). In contrast, WM associations were weak and inconsistent (Raw β = 0.2; harmonised β ≈ −0.2), with limited statistical significance across methods, likely reflecting cohort characteristics and residual scanner-related variability. Image-based harmonisation (SynthSR) yielded slightly attenuated effect sizes across several IDPs (e.g., Brain β = −0.3; GM β = −0.4), whereas statistical harmonisation approaches maintained comparable magnitudes to Raw estimates. These findings indicate that harmonisation preserved biologically plausible age– volume relationships and, in some cases, strengthened effect detectability without introducing directionally inconsistent associations.

**Figure 13.**
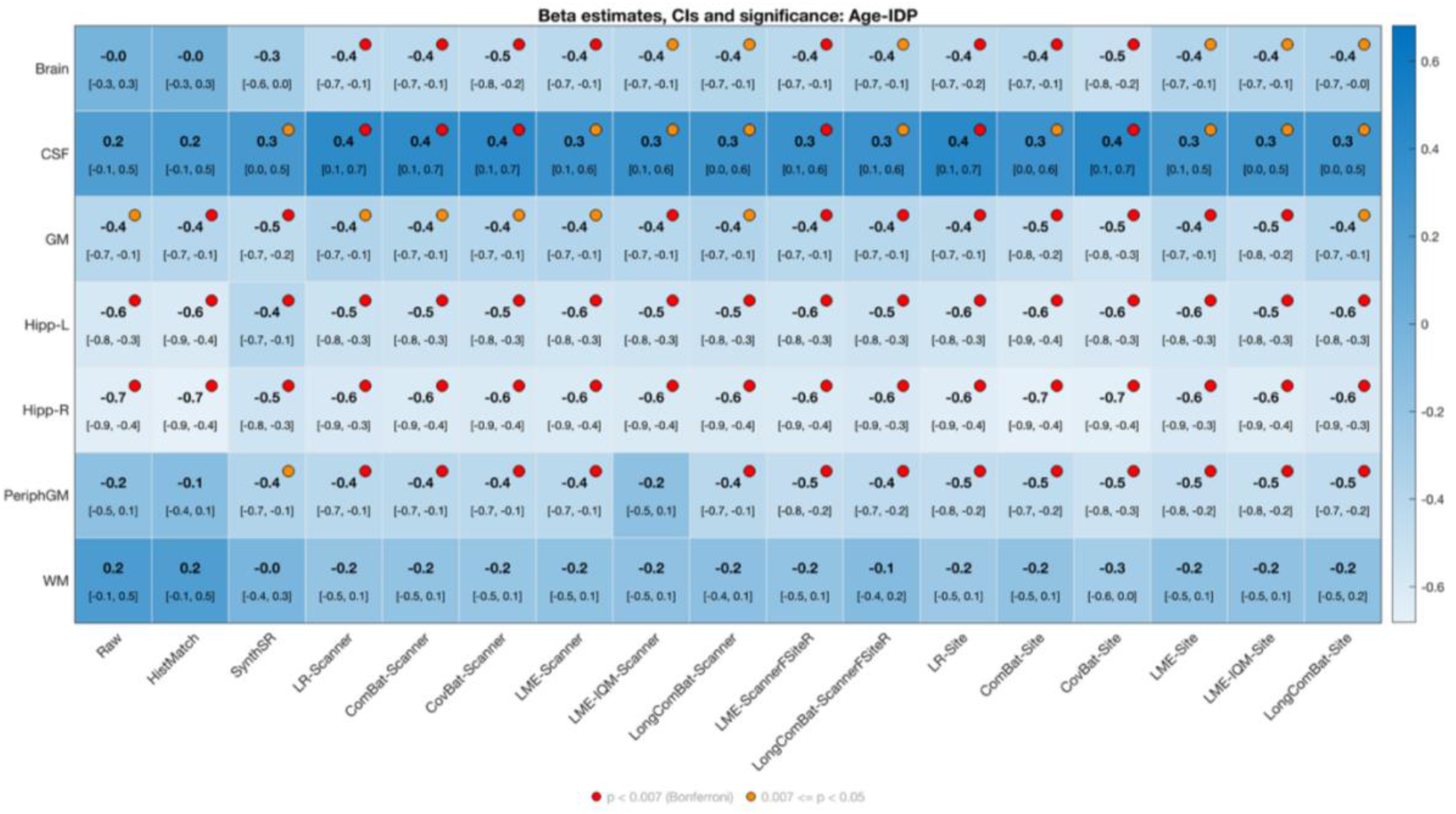
Age-related associations with raw and harmonised IDPs. Effect sizes (β estimates), 95% confidence intervals, and p-values for the fixed effect of age were estimated from linear mixed-effects evaluation models for each harmonisation method and IDP. The markers are highlighted with different colours to show the significance of *p*-values – red *p*<0.007 (Bonferroni corrected significant), orange 0.007<*p*<0.05 (uncorrected significant), grey *p*>0.05 (not significant)

##### Association between IDPs and timepoint

**Figure 14** shows timepoint-related effects on IDPs estimated using linear mixed-effects models in the test–retest data before and after harmonisation (Also see supplementary excel sheet ‘*biological_variability_Time_IDP*’ for more details).

**Figure 14.**
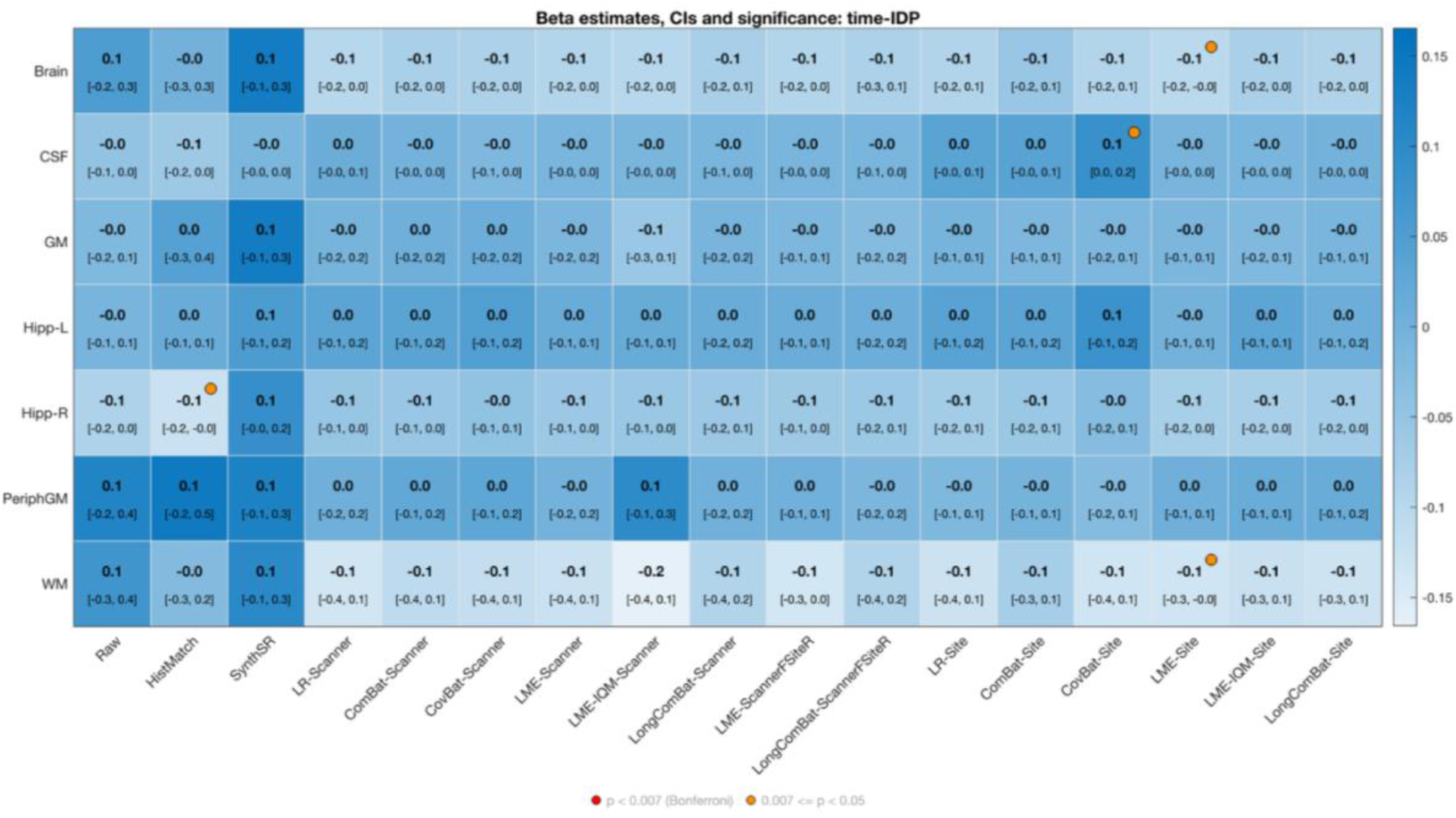
Timepoint-related associations with raw and harmonised IDPs. Effect sizes (β estimates), 95% confidence intervals, and p-values for the fixed effect of age were estimated from linear mixed-effects evaluation models for each harmonisation method and IDP. The markers are highlighted with different colours to show the significance and non-signifiance of I-values – red *p*<0.007 (Bonferroni corrected significant), orange 0.007<*p*<0.05 (uncorrected significant), grey *p*>0.05 (not significant)

Given the short inter-scan interval (∼1 year), time–IDP associations were expected to be modest, and this was reflected across harmonisation strategies. For all IDPs, effect sizes were small (typically |β| ≤ 0.1) and confidence intervals largely included zero. Whole-brain estimates ranged from β = 0.1 (Raw) to approximately −0.1 under most harmonised models, without consistent statistical significance. GM and hippocampal volumes showed near-null associations (β ≈ 0.0 to −0.1) across methods. CSF and peripheral GM demonstrated similarly small effects (|β| ≤ 0.1), with only isolated nominal findings (p < 0.05) under select site-based models. WM estimates ranged from β = 0.1 (Raw) to approximately −0.2 in some scanner-level models but remained small in magnitude and largely non-significant. No harmonisation method introduced systematic longitudinal trends or widespread significant time effects, indicating preservation of expected test–retest stability.

### 3.3 IQM importance in LME-IQM harmonisation

#### 3.3.1 IQMs capturing site effect (LME-IQM-Site)

**Figure 15** summarises the PCA-IQM components selected for additive and multiplicative correction in site-based harmonisation across IDPs (also see supplementary excel sheet – ‘*IQM-harmonisation_IQMImportance*’ for important QCs and their loadings in PCA). Ten PCA-IQM components were consistently identified across IDPs, indicating a shared QC-related structure underlying site effect. Both additive and multiplicative correction selected the same set of PCA-IQM components (except PCA-IQM11), with IDP-dependent component selection patterns. Loadings of the selected PCA-IQM components demonstrated that site-related variability was captured by a small number of interpretable image-quality dimensions. Across both additive and multiplicative correction, consistently selected components reflected (i) tissue intensity dispersion and scaling (e.g., SDIntensity_CSF/WM, MEAN_dsmri), (ii) spatial resolution and smoothness (FWHM measures, voxel spacing), (iii) contrast-to-noise structure (SNR-derived metrics, wm2max, FBER), (iv) bias-field and intensity non-uniformity (biasICR, noiseNCR), and (v) partial volume and segmentation stability (rpve_gm/csf, resolutionRMS). The recurrence of these components across IDPs indicates that site effects operate through structured, low-dimensional variations in contrast, smoothness, noise characteristics, and intensity scaling rather than arbitrary scanner offsets. Importantly, additive and multiplicative corrections converged on largely overlapping PCA-IQM components, suggesting that both location and scale biases arise from shared underlying image-quality mechanisms. Components such as PCA-IQM3 and PCA-IQM7, selected across nearly all IDPs, were dominated by smoothness, CSF intensity distribution, and resolution-related metrics, highlighting reconstruction- and contrast-driven site differences.

**Figure 15.**
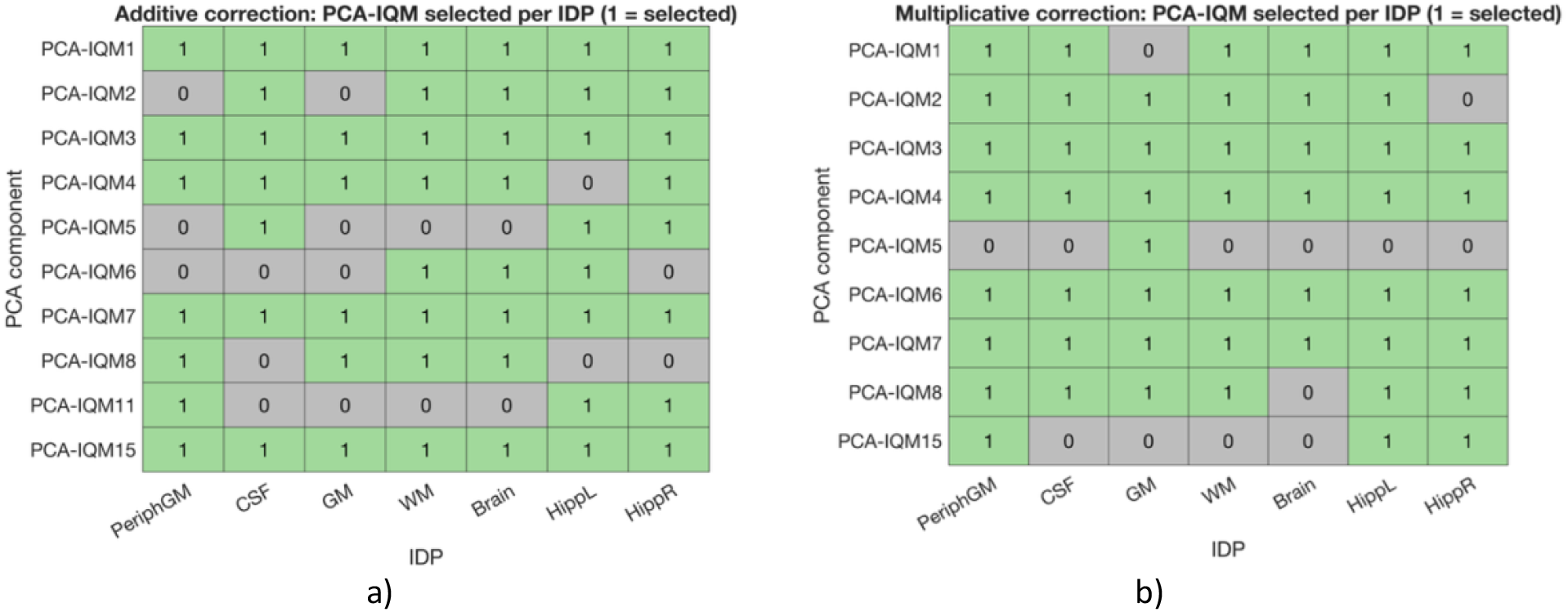
PCA-IQM components selected for site-based batch correction using LME-IQM-Site method – a) components selected for additive batch correction a) components selected for multiplicative batch correction.

#### 3.3.2 IQMs capturing scanner effect (LME-IQM-Scanner)

**Figure 16** summarises the PCA-IQM components selected for additive and multiplicative correction in scanner-based harmonisation across IDPs (also see supplementary excel sheet – ‘*IQM-harmonisation_IQMImportance*’ for important QCs and their loadings in PCA). For scanner-based harmonisation, a more restricted subset of PCA-IQM components was selected compared to site-based correction. Across IDPs, multiplicative correction consistently selected PCA-IQM7 and PCA-IQM9, with PCA-IQM6 also selected for the majority of IDPs, whereas PCA-IQM1 was only selected for WM and Hipp-R. Additive correction was comparatively sparse, with limited component selection and primarily restricted to PCA-IQM1, PCA-IQM7 and PCA-IQM9 in selected IDPs. Inspection of PCA loadings indicated that these scanner-associated components were dominated by IQMs reflecting tissue intensity dispersion (SDIntensity_CSF/WM), intensity dynamic range (WM percentiles, weightedIQR), contrast scaling (wm2max), partial volume measures (rpve_gm), noise structure (noiseNCR), bias-field inhomogeneity (biasICR), and effective spatial resolution (resolutionRMS). In contrast to the broader QC structure observed for site effects, scanner-related variability was largely captured by a small number of components centred on contrast, resolution, and intensity scaling properties. Notably, components associated with spatial smoothness and contrast structure (e.g., PCA-IQM7) were selected across nearly all IDPs under multiplicative correction, whereas additive effects were limited and IDP-dependent. This pattern indicates that scanner-related variability was captured by a smaller subset of QC dimensions and was more frequently associated with contrast, intensity dispersion, and resolution-related metrics.

**Figure 16.**
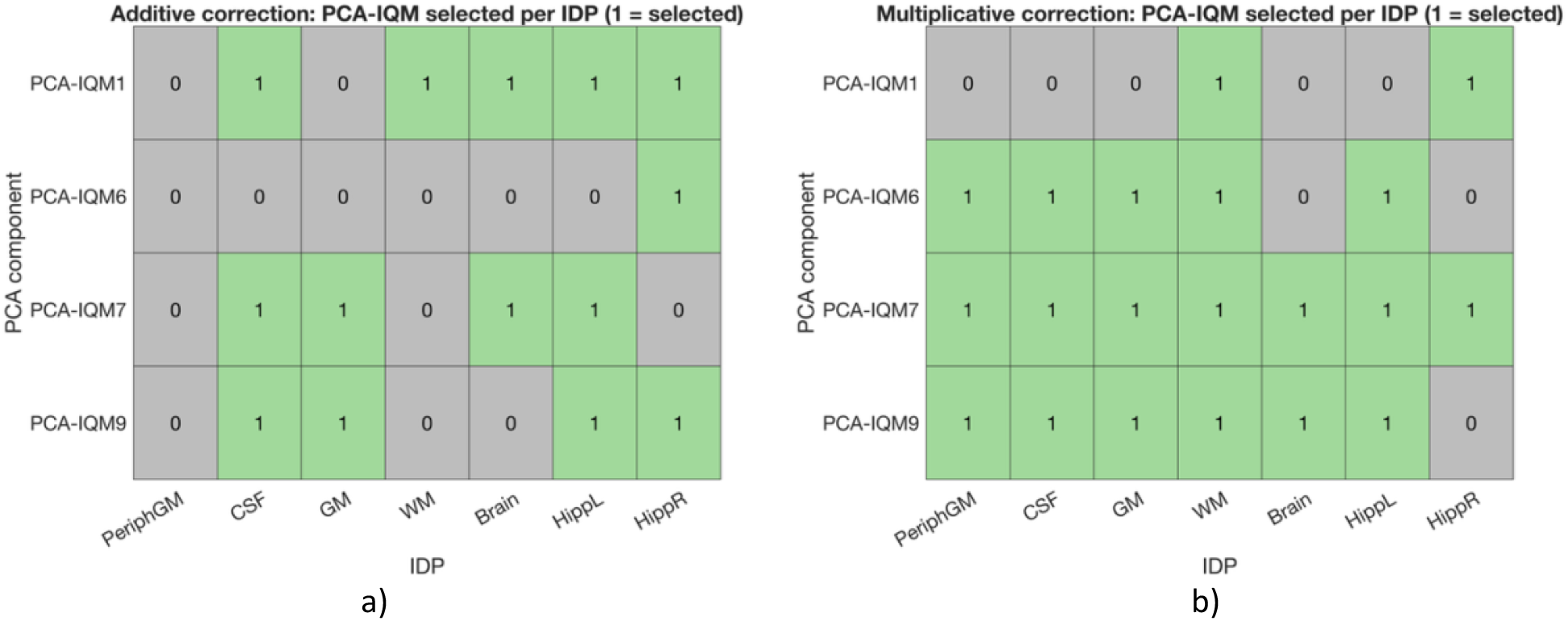
PCA-IQM components selected for scanner-based batch correction using LME-IQM-Scanner method – a) components selected for additive batch correction a) components selected for multiplicative batch correction.

## 4. Discussion

This Dementias Platform UK (DPUK) network study represents the first comprehensive evaluation of repeatability and reproducibility of volumetric MRI measures acquired using harmonised PET/MR imaging protocols across eight sites. Using a multisite, multiscanner test–retest dataset, we systematically characterised IDP variability across repeatability, intra-scanner, and inter-scanner acquisition settings. Within-subject variability analysis across categories showed that repeatability was high (i.e. variability was low) for all volumetric IDPs, with modest increases in variability under intra-scanner conditions, and substantially greater variability observed for some measures in inter-scanner settings. We evaluated a diverse set of harmonisation (image-based and statistical) approaches to mitigate site- and scanner-related batch effects. Our findings showed that statistical methods explicitly modelling site effects and repeated-measure structure in this data provided the most consistent balance between reducing technical variability and preserving biological associations.

Insights from multisite test–retest datasets such as this provide valuable information for the design and optimisation of clinical trials including neuroimaging measures. The median hippocampal variability observed in this study is consistent with previously reported reproducibility estimates for FSL-FIRST segmentation. In particular, the raw repeatability values (≈2.5% left, ≈2.7% right) closely match previously reported back-to-back reproducibility estimates. For example, median differences of approximately 2.5–2.7% (Cover et al., 2016) and 2.8–3.6% (Mulder et al., 2014) have been reported for the left and right hippocampus, respectively, indicating that the magnitude of measurement variability in our dataset is comparable to established benchmarks. Interpreted within this framework, the median RPD values in our work represent the effective measurement variability expected in a clinical trial setting and can therefore be used to estimate the reproducibility of candidate analysis pipelines, informing prospective selection of processing strategies and sample size requirements.

As a potential illustrative application, following the square-root-of-N rule described in (Cover et al., 2016), relative sample size can be estimated as the square of the ratio of variability between conditions. Using the left hippocampus as a representative example, intra-scanner variability in the raw data (3.2%) corresponds to a relative group size of 1.64 (=(3.2/2.5)^2^), while inter-scanner variability (3.5%) corresponds to 1.96(=(3.5/2.5)^2^) compared to repeatability. In practical terms, if a study designed under repeatability conditions required 10 subjects per group to detect a given atrophy rate, the same study would typically require approximately 16 subjects under intra-scanner conditions and 20 subjects under inter-scanner conditions. Although the present dataset consists of healthy participants, these relative differences in variability are expected to translate to longitudinal studies of neurodegeneration, where hippocampal atrophy could be a key endpoint.

Harmonisation reduces this excess variability and therefore the associated sample size penalty. For example, for the left hippocampus, intra-scanner variability can be reduced from 3.2% to approximately 1.9%, and inter-scanner variability from 3.5% to approximately 2.6%. In practical terms, this means that a study requiring 16 subjects under raw intra-scanner conditions may require substantially fewer subjects after harmonisation, while an inter-scanner study requiring ∼20 subjects may be reduced to close to the repeatability-level requirement. These gains, however, depend on the baseline level of variability. In repeatability and intra-scanner settings, where variability is already low (e.g. hippocampal RPD ≈ 2.5–3%), most harmonisation approaches produce results comparable to raw measurements, and additional correction offers limited benefit. In contrast, inter-scanner heterogeneity introduces substantially greater variability for several IDPs, particularly white matter and CSF volumes (raw RPD ≈ 13% and ≈ 19%, respectively), where harmonisation plays a more critical role in reducing variability and improving sensitivity to longitudinal effects. In addition to reducing variability, harmonisation also influenced the consistency of subject-level measurements across timepoints. The repeatability group showed high rank-order consistency across all IDPs, while intra- and particularly inter-scanner conditions introduced greater variability in subject ranking for some measures (notably white matter, brain volume, and peripheral grey matter). For key clinical measures such as hippocampal and CSF volumes, rank-order consistency remained high across all categories, indicating that relative differences between individuals are largely preserved even without harmonisation. Where ranking consistency was reduced, harmonisation improved stability across timepoints, suggesting that it can mitigate noise-driven reordering effects. This is particularly relevant for clinical trials, as preservation of subject ordering indicates that measurement noise does not distort inter-individual differences in longitudinal change, thereby supporting robust detection of treatment effects. Taken together, these findings indicate that harmonisation is most beneficial when acquisition-related variability exceeds the repeatability noise floor and can improve both the statistical efficiency and interpretability of multisite clinical studies.

When data were pooled across test-retest categories, additive (mean-shift) site-related batch effects assessed via omnibus model comparison emerged as the dominant source of technical bias. The persistence of these additive site effects after several harmonisation methods, particularly those with scanner-only batch effect removal strategy, indicates that correcting scanner-related differences alone is insufficient when site-level variability remains unmodelled. Approaches that explicitly incorporated site effects and repeated-measure structure were more effective at reducing these global mean shifts (LME, IQMs method, longitudinal ComBat), underscoring the importance of aligning batch specification with the dominant source of technical variation. Even when omnibus tests indicated no significant additive batch effect, pairwise site contrasts and multivariate site-wise deviations often revealed residual batch differences for specific sites. This demonstrates that reductions in global mean shifts in measurements can coexist with localized site-specific heterogeneity, and that harmonisation success cannot be inferred from a single omnibus statistic alone. This has direct implications for downstream analyses, where such residual structure could bias group comparisons or inflate confidence in harmonisation effectiveness if not explicitly examined. In addition, the relative absence of significant multiplicative (scaling) batch effects across most IDPs suggests that variance-related differences between sites were not a major driver of technical heterogeneity in this dataset.

The biological associations (between-subject variability, age-IDP, timepoint-IDP) showed important insights into how different harmonisation strategies balance the trade-off between attenuating technical variability and preserving or revealing true biological associations. Across most IDPs, between-subject variability was already high prior to harmonisation and remained largely unchanged across methods, indicating that harmonisation did not substantially compress inter-individual differences. A notable exception was white matter volume, where raw ICC values were comparatively low; harmonisation approaches (especially methods that modelled site effects and repeated-measure structure) improved ICC substantially, suggesting that removal of site-related technical variance allowed biologically meaningful variability to emerge more clearly. Age associations with hippocampus volumes emerged as the most robust and reliable biological signal across analyses, both before and after harmonisation. This consistency reinforces the suitability of hippocampus measures as biologically sensitive markers in aging and dementia studies and provides a useful reference for assessing whether harmonisation preserves expected effects. In contrast, age associations for other IDPs were weaker and less precisely estimated prior to harmonisation, highlighting the challenge of detecting subtle biological effects in small, heterogeneous clinical datasets. The timepoint-IDP association analysis, included here as a negative control in the test–retest setting, further supports this interpretation. Across harmonisation strategies, time effects remained small and statistically non-significant, indicating that harmonisation did not introduce spurious longitudinal trends. Together, these findings suggest that well-performing harmonisation approaches can improve the precision of biological effect estimation while preserving expected signal structure and between-subject variability.

Taken together, the results support a clear hierarchy of harmonisation performance. In image-based methods histogram matching consistently showed unstable behaviour across evaluation metrics and was unsuitable for harmonising batch structure in this dataset. SynthSR demonstrated strong overall performance, reducing batch variability in some IDPs and preserving biological variability, but requires cautious interpretation given its tendency to reduce between-subject differences and its potential to differentially affect regionally specific measures in longitudinal settings (for, e.g., increased within-subject variability in inter-scanner group and significant additive batch effect in hippocampus). Among statistical approaches, scanner-only batch correction methods that do not account for repeated measures overall underperformed, mixed batch formulations (i.e. scanner as fixed effect and site as random effect) provided a robust compromise, and explicit site-based harmonisation was generally more effective in pooled analyses. Among approaches using site as batch, linear mixed-effects and IQM-informed models showed the most consistent performance across evaluation domains, balancing technical correction with preservation of biological variability and temporal stability. In this dataset, variants of linear regression, ComBat, and CovBat methods performed comparably with each other across several evaluation metrics, but their performance was limited when metrics sensitive to subject-level and repeated measures structure were considered. Despite these methods being mainly intended for harmonising cross-sectional datasets, we included them given their widespread adoption in the neuroimaging literature and allowing us to empirically assess how well such commonly applied methods generalise to a small, clinically realistic dataset. Longitudinal ComBat which implicitly accounts for repeated-measure structure showed improved performance in several evaluation metrics but also did not achieve consistent performance across all domains (e.g., within subject variability, multivariate batch differences), highlighting the sensitivity of ComBat-based approaches to sample size across batches and underlying model assumptions. Specifically, these methods require specifying batch identifiers in the model and need sufficient per-batch sample sizes to reliably estimate batch effects (minimum 20-30 samples per batch are recommended (Orlhac et al., 2022)). The need for discrete labels may oversimplify scanner- or site-related heterogeneity, which often varies continuously (e.g., across scanner upgrades, acquisition parameter differences, or gradual hardware drift), leaving residual confounds even after harmonisation. The dependence on group-level estimation further limits clinical translational potential to small sample sizes. By contrast, our image quality metrics-informed approach modelled scanner and site-related variability using continuous covariates within a mixed-effects framework rather than categorical batch labels. Our approach showed relatively stable behaviour across evaluation metrics and IDPs, suggesting increased flexibility in capturing and reducing heterogeneous technical effects. Importantly, this formulation also offers a potential pathway for harmonisation in settings where batch information is unavailable (e.g., due to anonymisation) or where sample sizes per batch are insufficient for stable estimation. Our findings highlight the potential of IQM-informed modelling as a flexible alternative to categorical batch definitions.

More broadly, these results emphasise that harmonisation should be viewed as a methodological step to consider in a study rather than a one-fits-all solution. Improvements in post-harmonisation variability or statistical alignment do not automatically imply recovery of the underlying biological signal, particularly when ground truth is unknown. The value of harmonisation depends on the downstream objective whether detecting longitudinal change, estimating treatment effects, or supporting predictive modelling. Controlled designs such as travelling-head studies, paired acquisitions, and test–retest datasets therefore play a critical role, providing a framework to quantify technical uncertainty and determine when harmonisation enhances, preserves, or potentially distorts clinically meaningful patterns. Establishing this comprehensive evaluation framework is essential for moving harmonisation from methodological optimisation toward reliable clinical utility.

## 5. Conclusion

Variability differences across acquisition settings have direct implications for the design and interpretation of multi-centre studies and clinical trials. Harmonisation can reduce this variability and bring multisite measurements closer to repeatability-level performance, while preserving subject-level ordering necessary for robust detection of treatment effects. In this context, harmonisation can substantially improve data comparability in multisite studies, but its benefits are method- and context-dependent. No approach was universally optimal across imaging-derived phenotypes or evaluation metrics, underscoring that harmonisation should not be treated as a one-size-fits-all solution. Our results highlight the value of a multi-metric evaluation framework in revealing both the benefits and unintended consequences of different strategies. By characterising the technical variability in their datasets, adopting the harmonisation approach that best addresses the type of effect, and evaluating success with multiple metrics, researchers can maximise the benefits of harmonisation while avoiding under- or over-correction, supporting more reliable use of clinical neuroimaging data in both research and translational settings.

## Supporting information

Appendix A - IQM based harmonisation

Supplementary Excel Sheet

## Data Availability

The DPUK PET/MR Network will make PET/MR imaging data and associated datasets available to the research community via controlled-access data-sharing procedures, in accordance with applicable data protection regulations.

https://github.com/gvbhalerao591/Harmonisation-Paper/

## 6. Code and data availability

- The code used to implement the harmonisation methods and evaluation metrics described in this manuscript is available at: https://github.com/gvbhalerao591/Harmonisation-Paper.
- In addition, these methods are provided as part of the *DiagnoseHarmonise* tool, with a Python implementation that enables researchers to apply the evaluation metrics to their own datasets: https://jake-turnbull.github.io/HarmonisationDiagnostics/
- The complete details on IQM-informed harmonisation method implementation are provided in *Appendix A* (https://github.com/gvbhalerao591/Harmonisation-Paper/tree/main/manuscript_materials).
- The DPUK PET/MR Network will make PET/MR imaging data and associated datasets available to the research community via controlled-access data-sharing procedures, in accordance with applicable data protection regulations.

## 7. Author Contributions

Gaurav Bhalerao - Conceptualisation, Methodology, Software, Validation, Formal analysis, Data curation, Writing - Original Draft, Visualization; Pawel Markiewicz - Investigation, Resources, Data curation; Jacob Turnbull - Software, Validation, Writing - Review & Editing; David Thomas – Methodology, Investigation, Resources, Writing - Review & Editing; Enrico de Vita - Resources, Writing - Review & Editing; Laura Parkes - Resources, Writing - Review & Editing; Gerard Thompson - Resources, Writing - Review & Editing; Jane MacKewn - Resources, Writing - Review & Editing; Georgios Krokos - Resources, Writing - Review & Editing; Catriona Wimberley - Resources, Writing - Review & Editing; William Hallett - Resources, Writing - Review & Editing; Li Su - Writing - Resources, Review & Editing; Paresh Malhotra - Resources, Writing - Review & Editing; Nigel Hoggard - Resources, Writing - Review & Editing; John-Paul Taylor - Resources, Writing - Review & Editing; Craig Ritchie - Resources, Writing - Review & Editing; Joanna Wardlaw - Resources, Writing - Review & Editing ; Paul Matthews - Resources, Writing - Review & Editing; Franklin Aigbirho - Resources, Writing - Review & Editing; John O’Brien - Resources, Writing - Review & Editing; Alexander Hammers - Resources, Writing - Review & Editing; Nick Fox - Resources, Writing - Review & Editing; Karl Herholz - Resources, Writing - Review & Editing; Frederik Barkhof - Resources, Writing - Review & Editing; Karla Miller - Resources, Writing - Review & Editing, Supervision, Funding acquisition; Julian Matthews - Resources, Writing - Review & Editing; Stephen Smith - Methodology, Resources, Writing - Review & Editing, Visualization, Supervision, Funding acquisition; Ludovica Griffanti - Conceptualization, Methodology, Writing - Review & Editing, Visualization, Supervision, Funding acquisition.

## 9. Acknowledgements

This work was supported by the UK Medical Research Council Dementias Platform UK (MR/T033371/1), Alzheimer’s Association Grant (AARF-21-846366), and supported by the NIHR Oxford Health Biomedical Research Centre (NIHR203316). The views expressed are those of the author(s) and not necessarily those of the NIHR or the Department of Health and Social Care. The Centre for Integrative Neuroimaging was supported by core funding from the Wellcome Trust (203139/Z/16/Z and 203139/A/16/Z). Author J-P.T is supported by the NIHR Newcastle Biomedical Research Centre (BRC). Author FB is supported by the NIHR biomedical research centre at UCLH. DT is supported by the UCLH NIHR Biomedical Research Centre. We would also like to thank the OxCIN Harmonisation Working Group for their valuable comments and discussions in the progression of this work.

## 10. Supplementary Materials

Supplementary material for this article is available: https://github.com/gvbhalerao591/Harmonisation-Paper/tree/main/manuscript_materials

