## Supplementary document for "Harmonising Structural Brain MRI from Multiple Sites with Limited Sample Sizes"

### Within-subject variability (RPD) for different imaging-derived phenotypes

a) Peripheral GM volumes

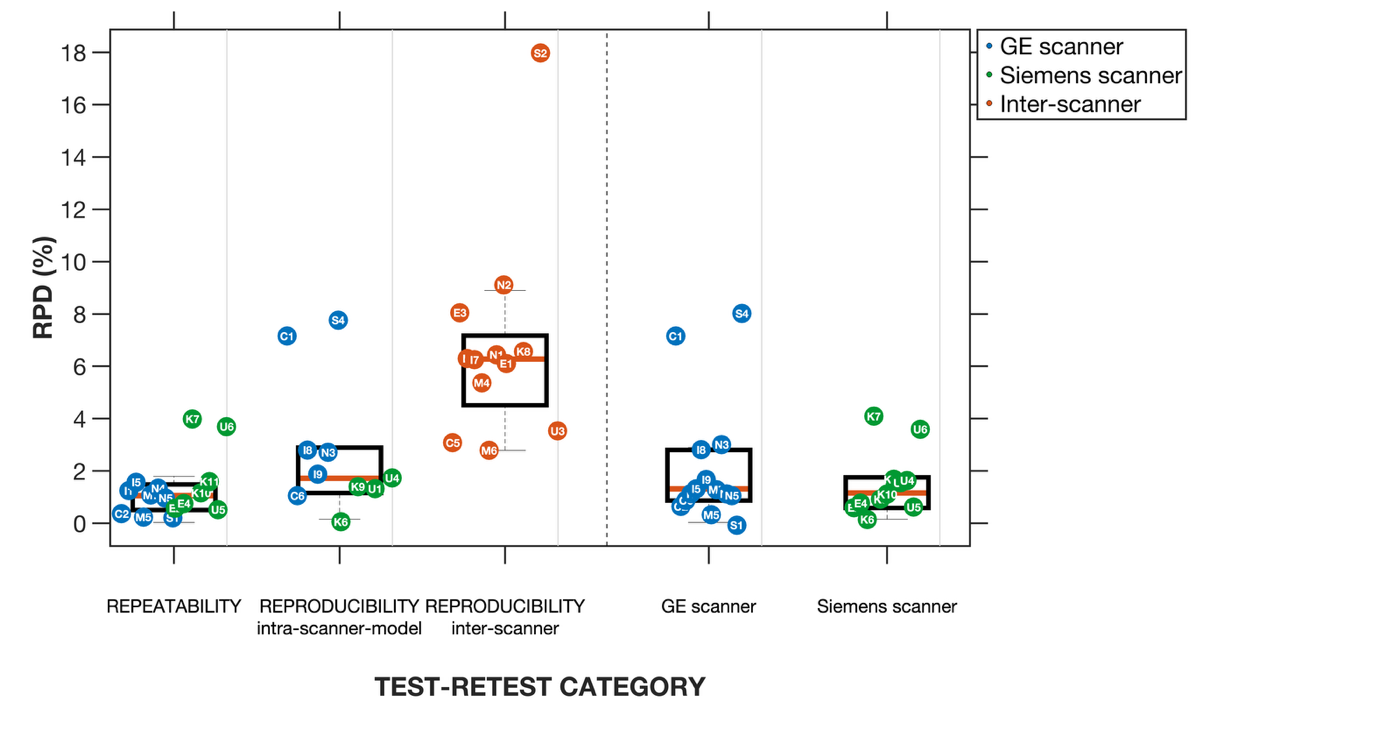

b) CSF volumes

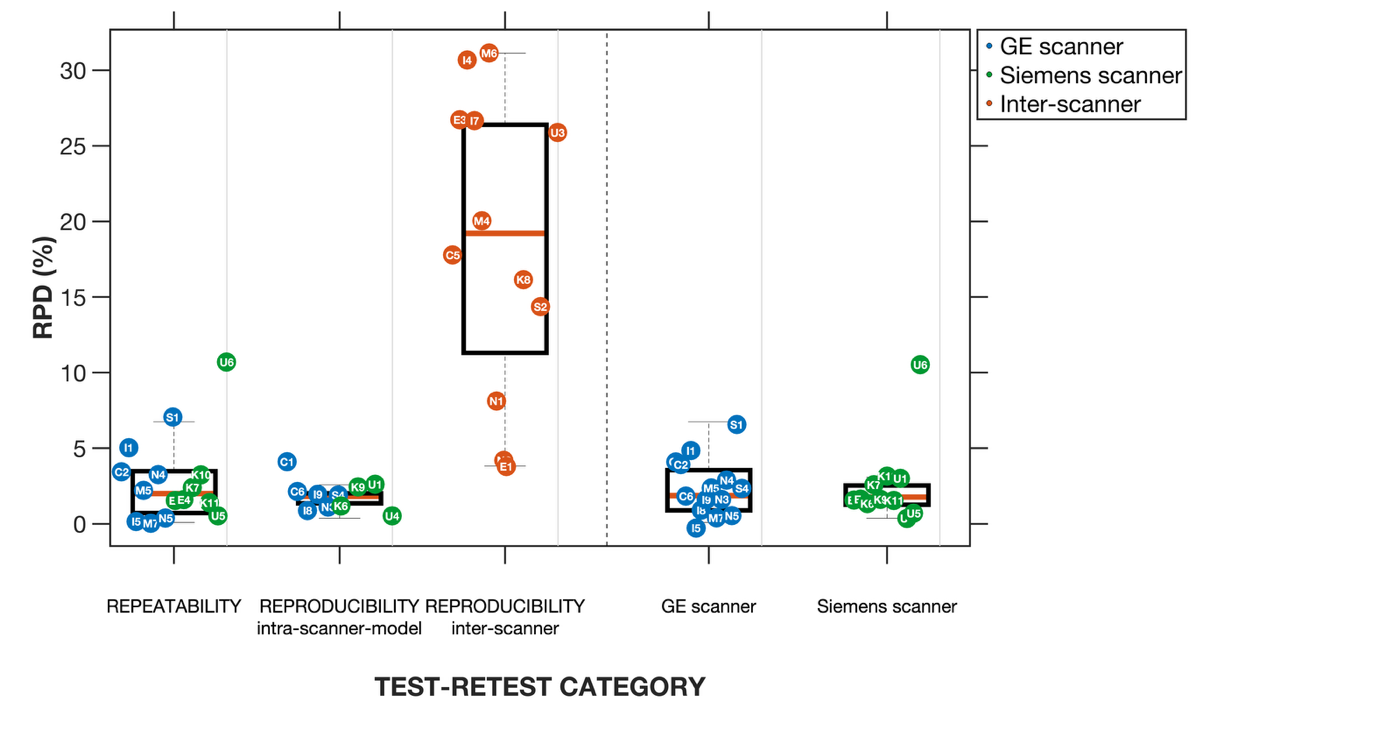

c) White Matter volumes

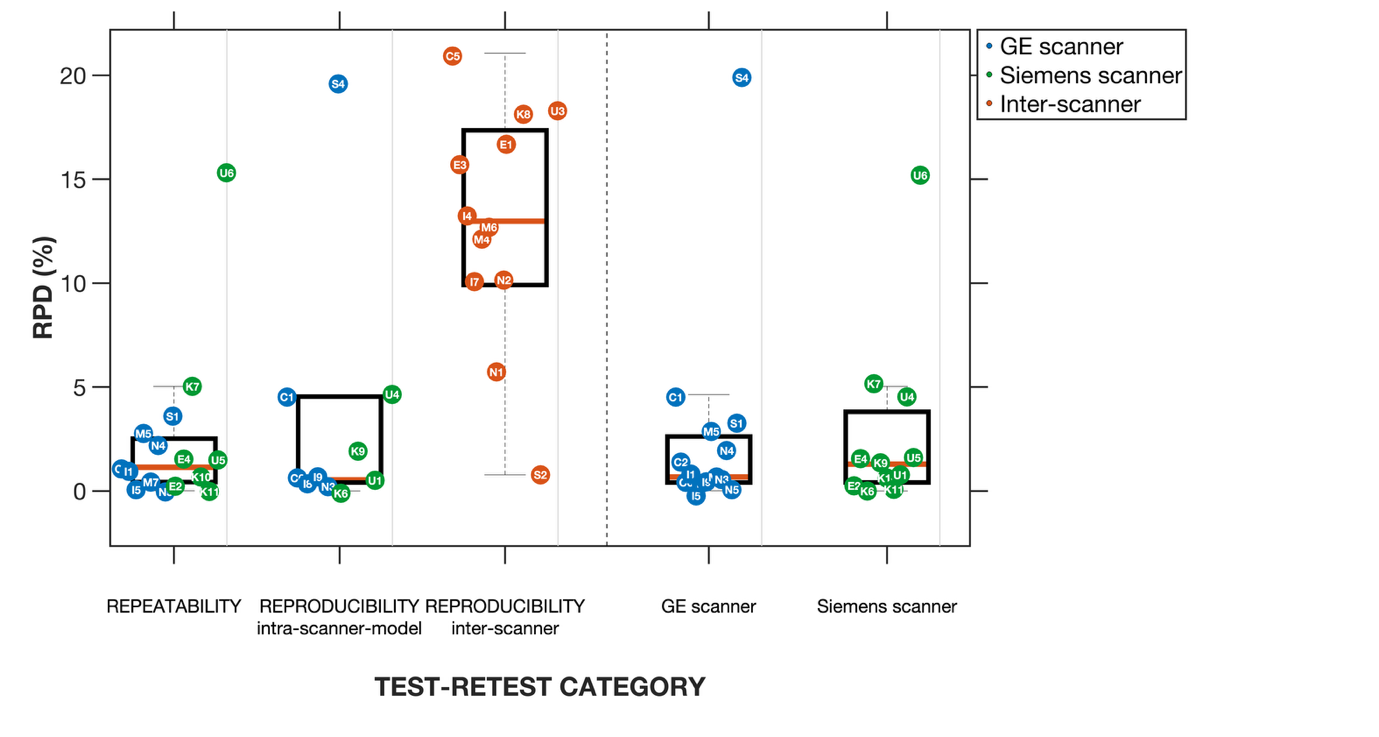

d) Gray matter volumes

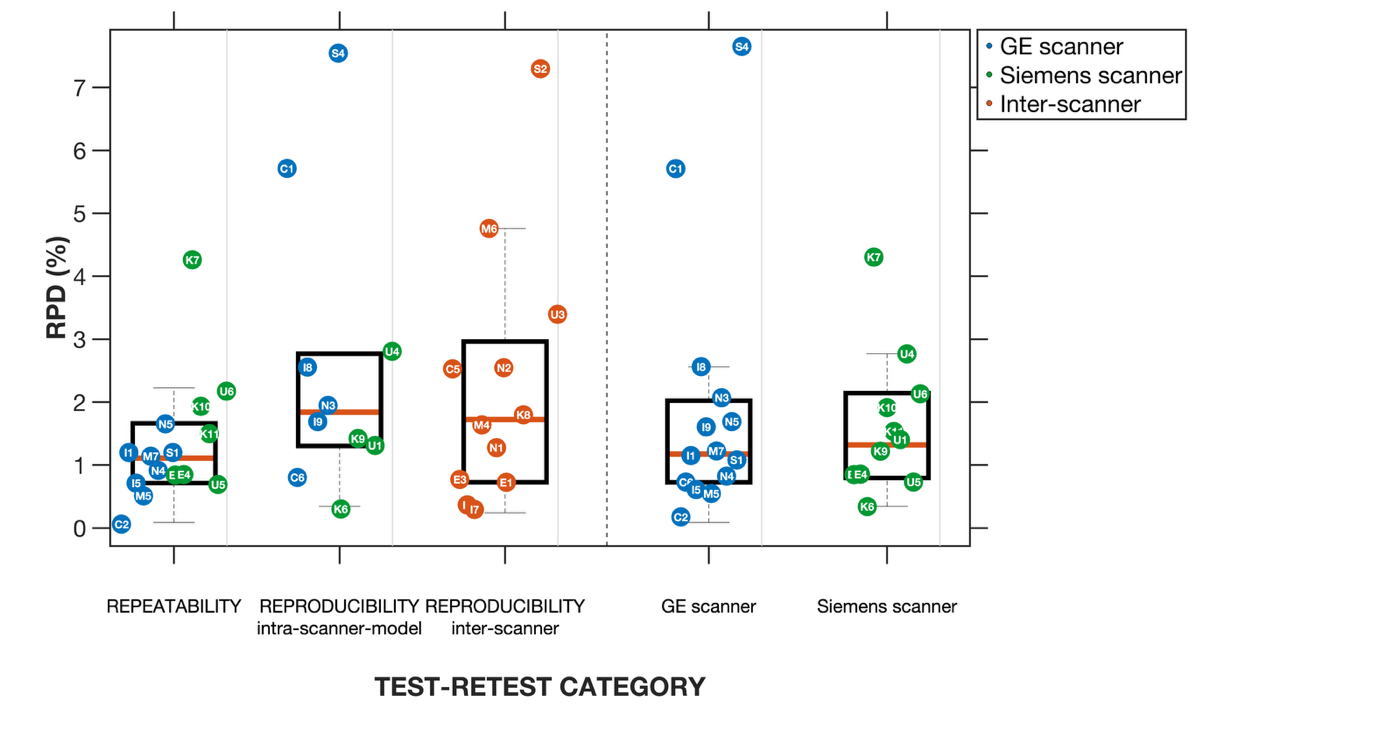

e) Left hippocampus volumes

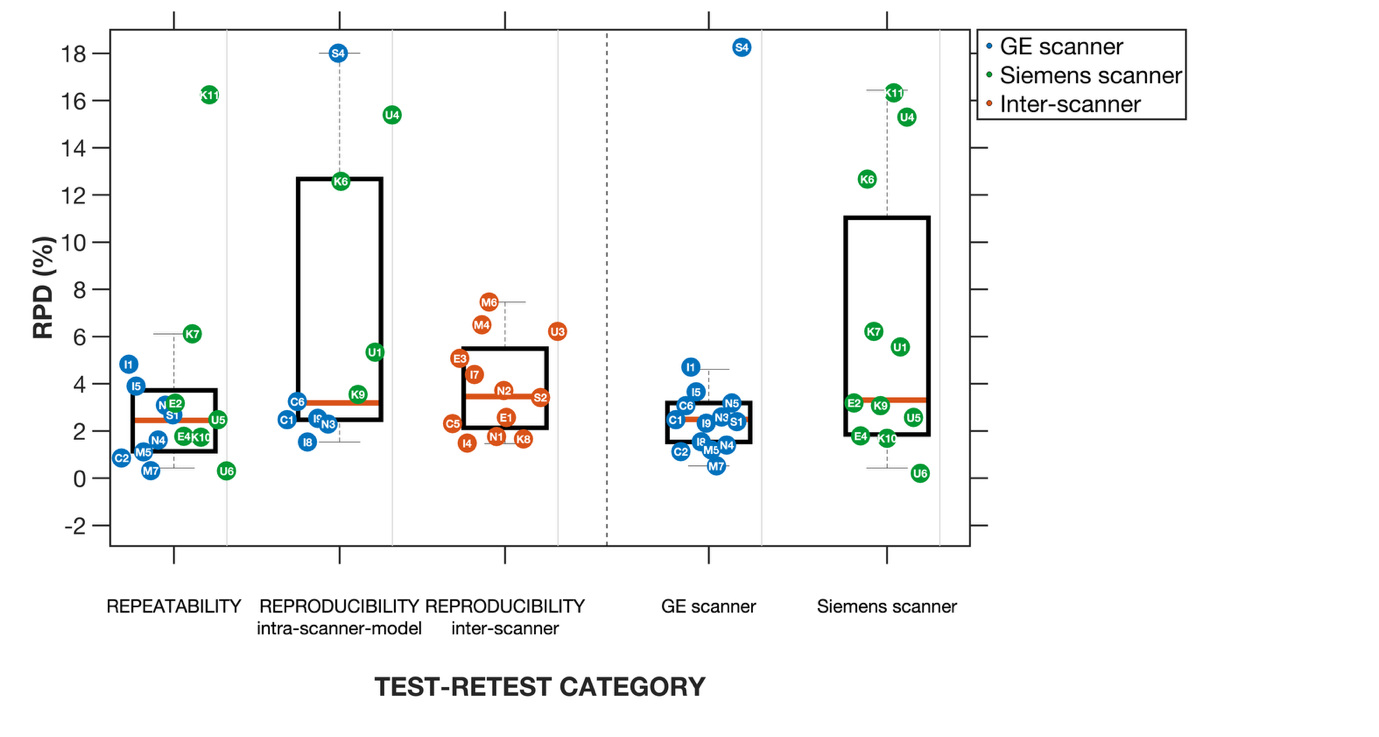

f) Right hippocampus

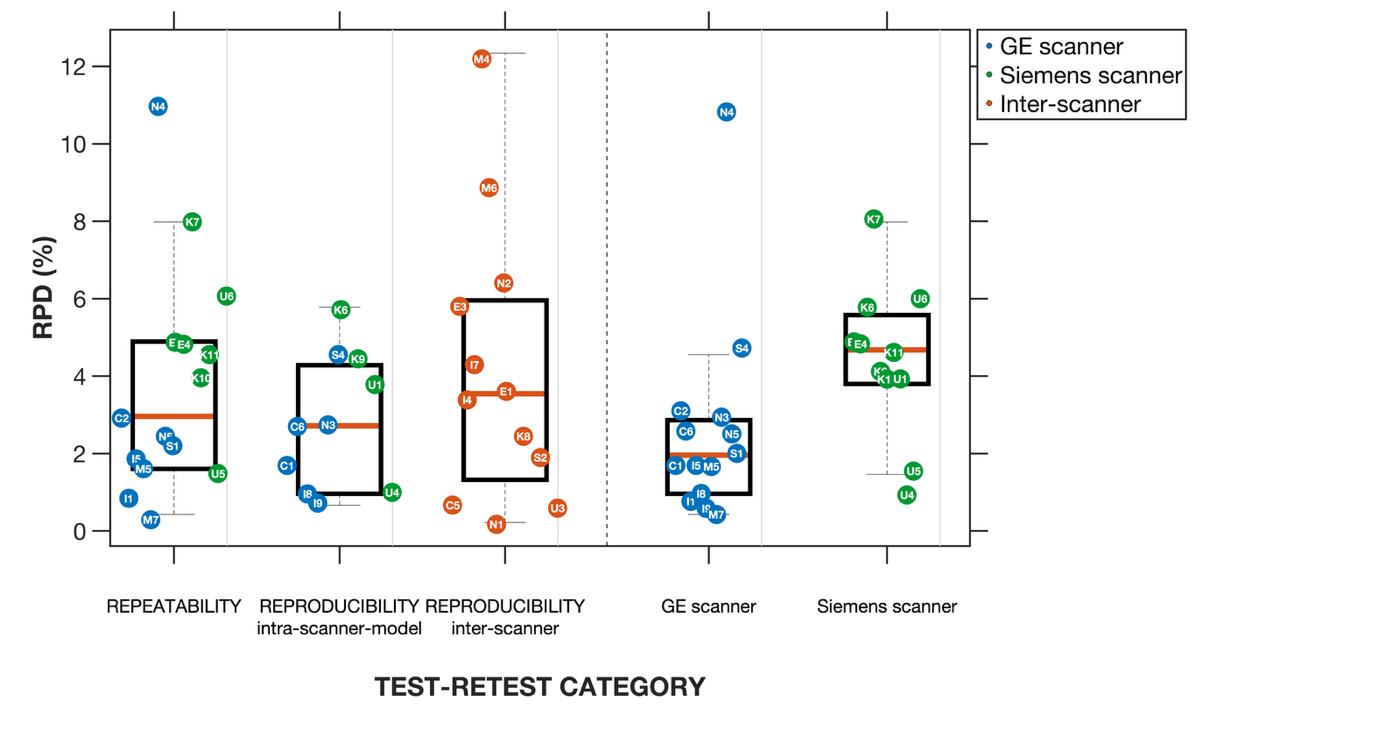

**Figure S1**. Within-subject IDP differences and visualisation of an outlier scan. Panel a) shows the relative percent difference (RPD, %) in IDPs panels a) to f) between two timepoints across test–retest categories: repeatability (same scanner and site), intra–scanner-model reproducibility (same scanner model, different site), and inter-scanner reproducibility (different scanner and site). Intra–scanner-model results (same or different site) are shown separately for GE and Siemens scanners.

### IDP distribution before (Raw) and after applying different harmonisation methods

a) Peripheral GM volumes

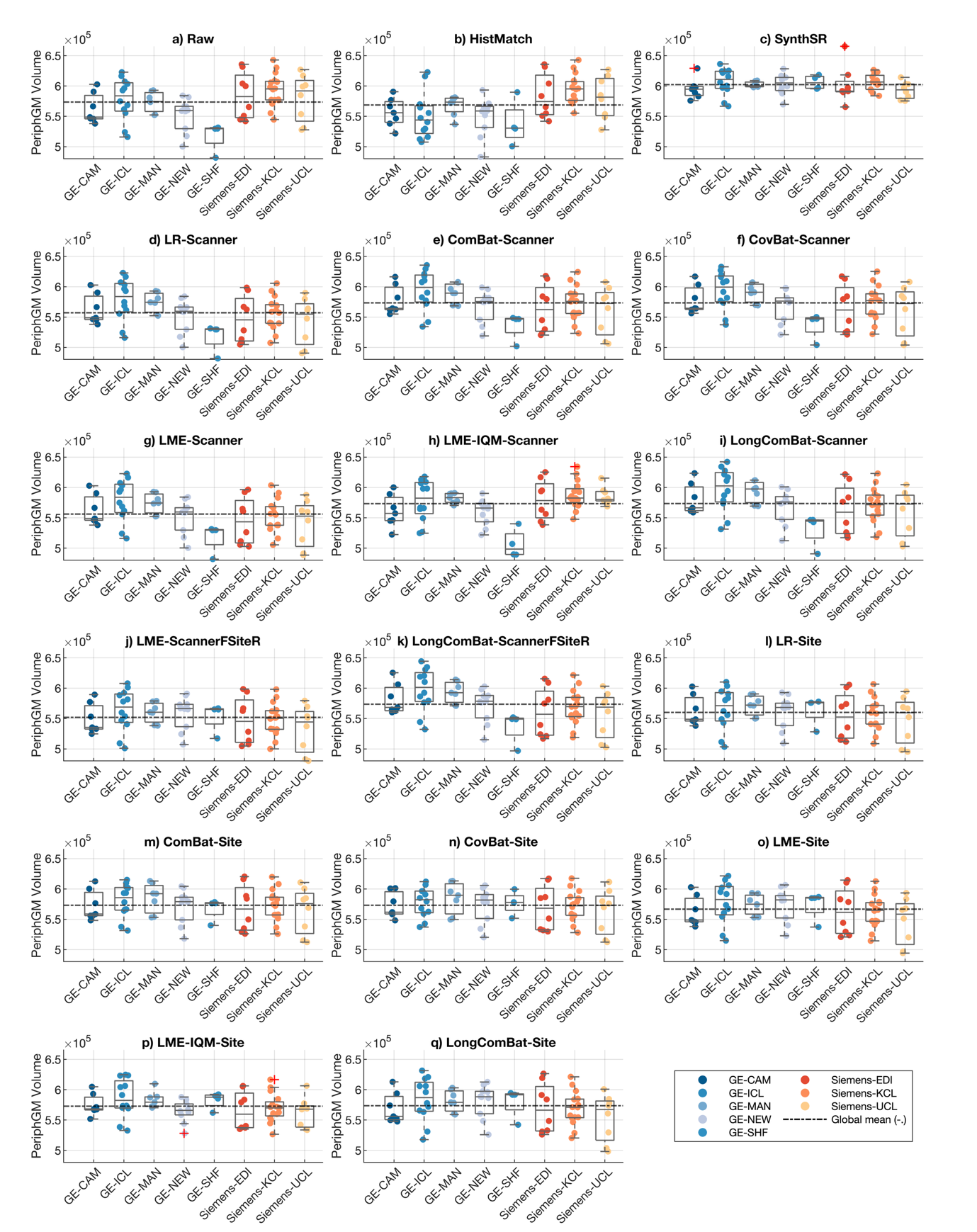

b) CSF volumes

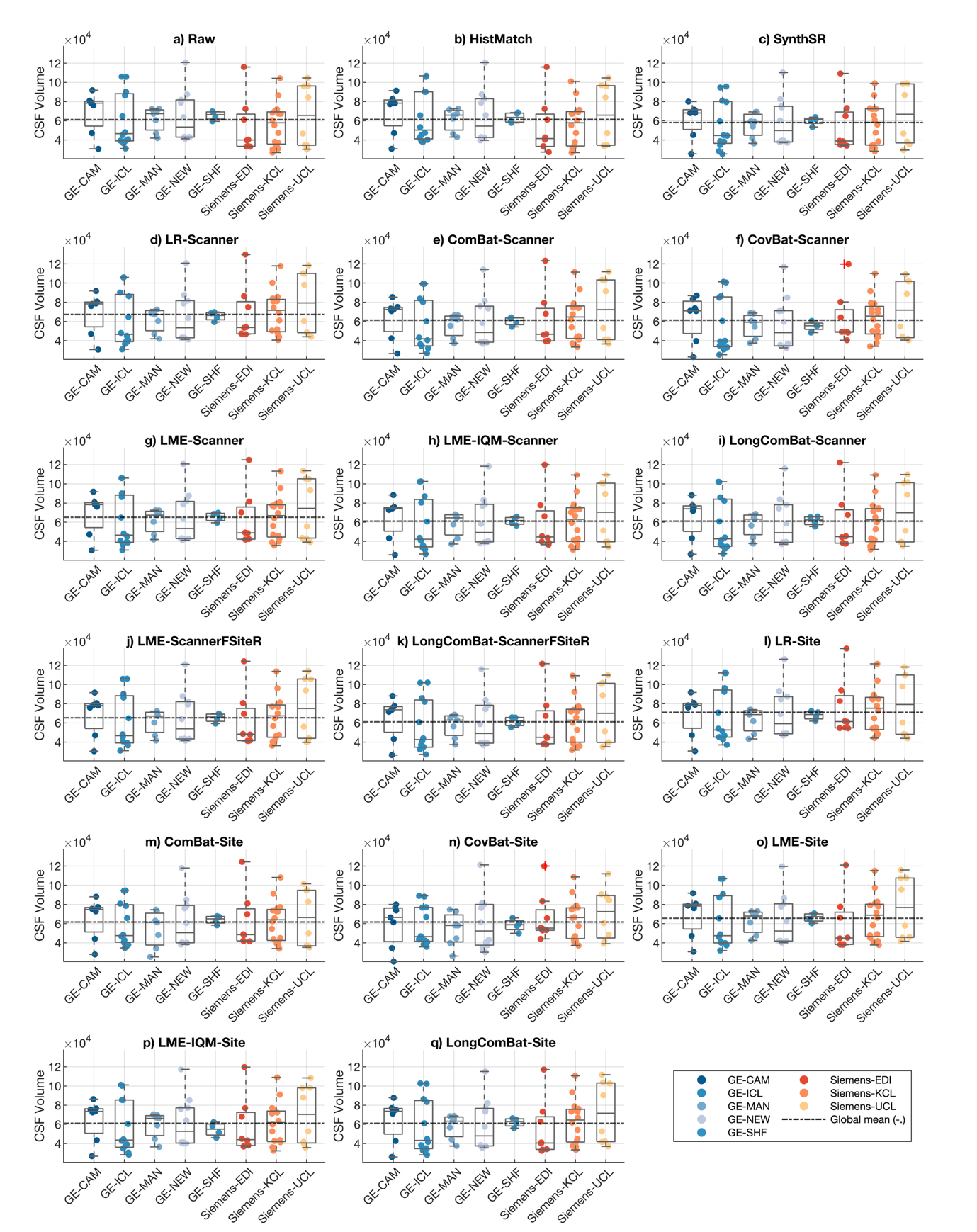

c) White matter volumes

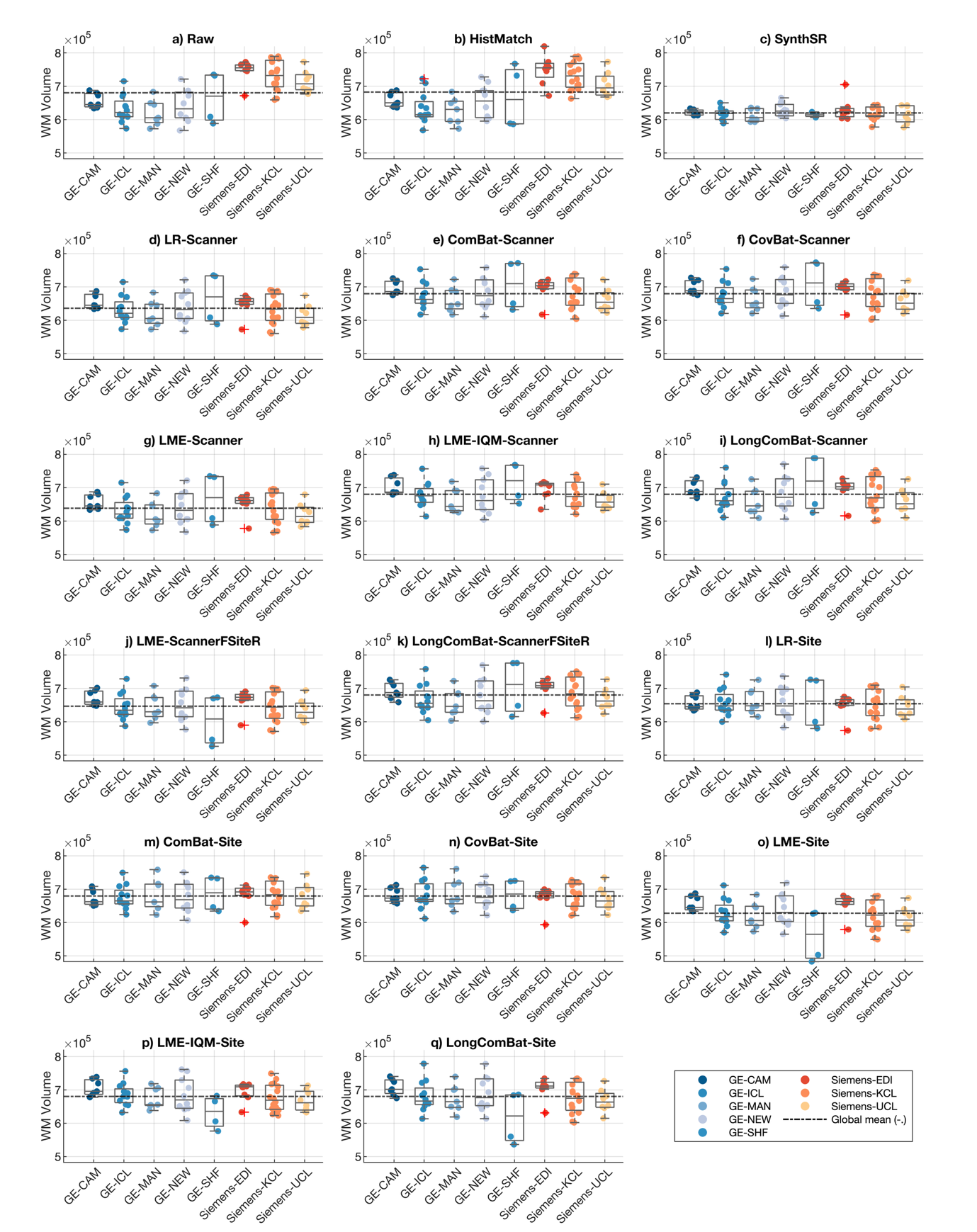

d) Gray matter volumes

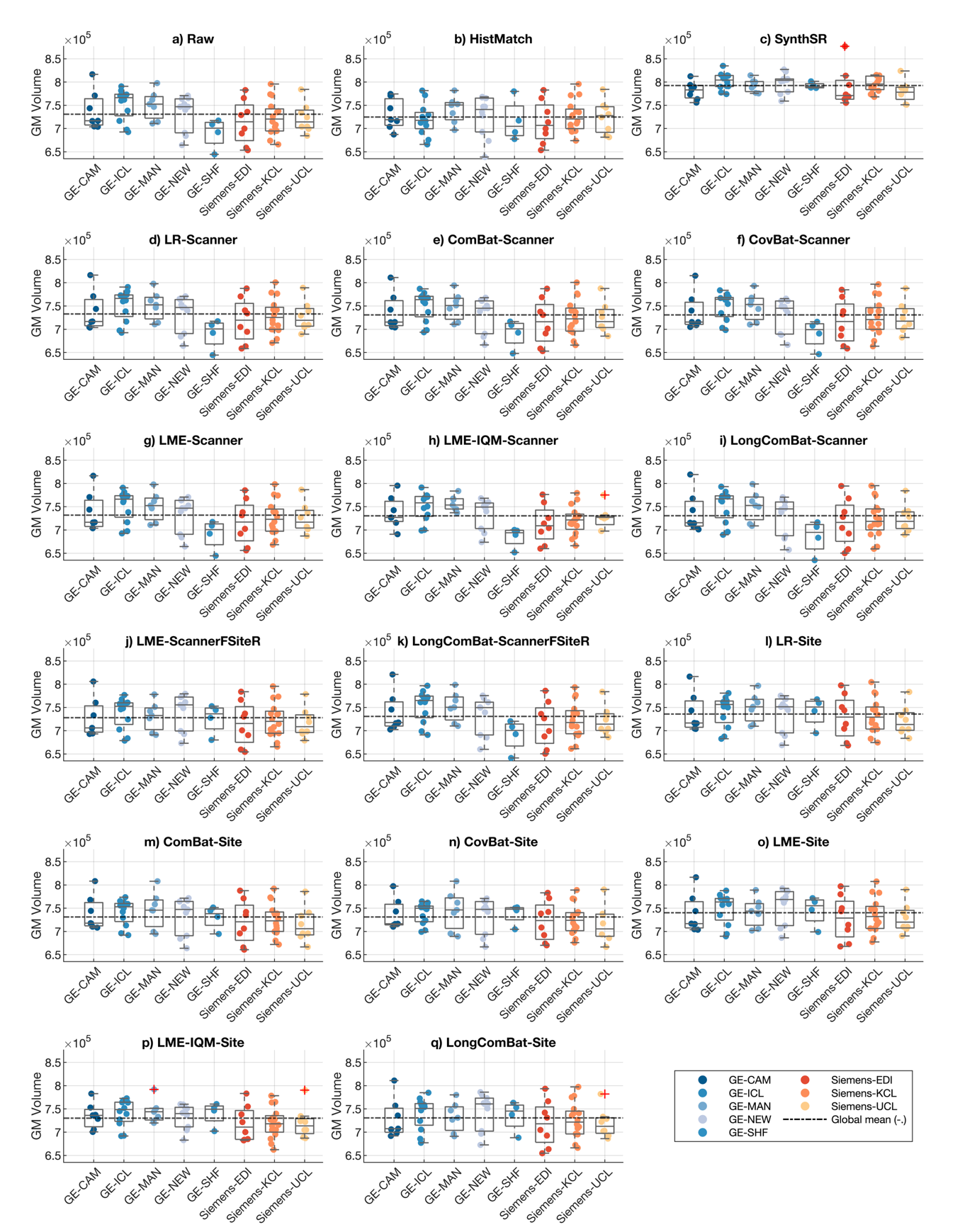

e) Left hippocampus volumes

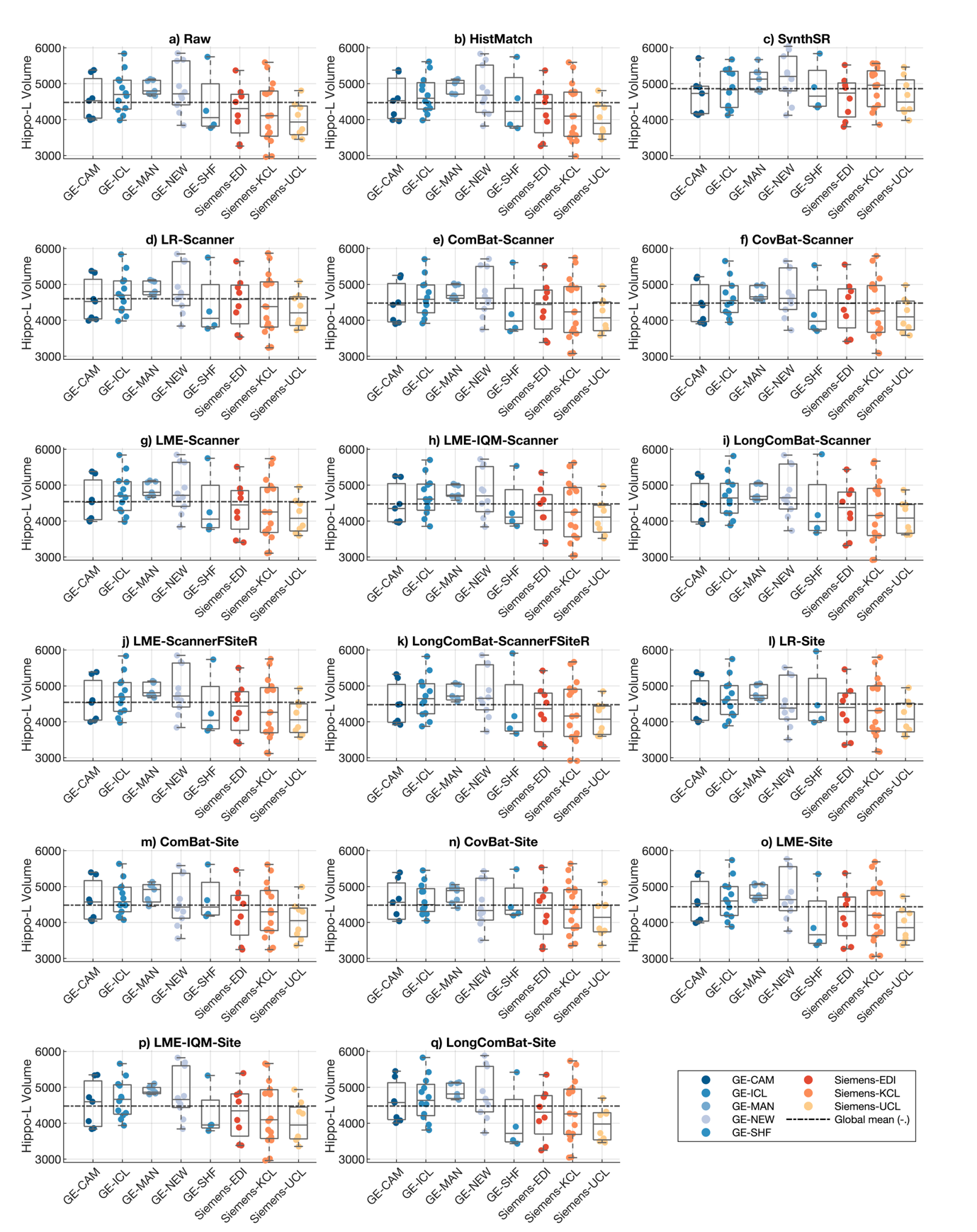

f) Right hippocampus volumes

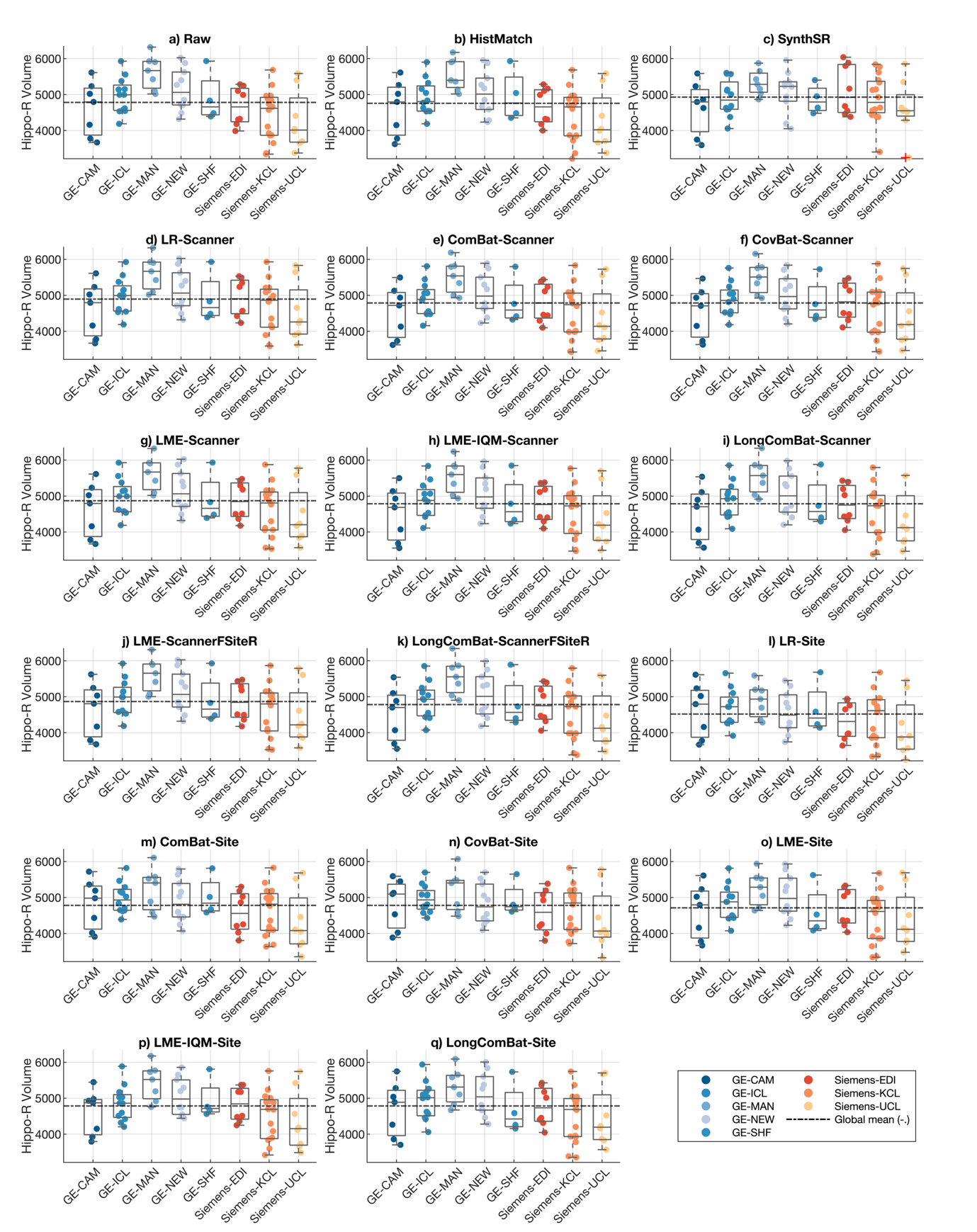

Figure S2. Distribution of IDPs across different sites and scanners before and after harmonisation
