## Appendix A - IQM based harmonisation for "Harmonising Structural Brain MRI from Multiple Sites with Limited Sample Sizes"

### 1 IQM-based Harmonisation

This appendix provides a complete technical description of the IQM-based harmonisation framework used in this study.

#### 1.1 Image Quality Metrics and Dimensionality Reduction

For each T1-weighted scan, image quality metrics (IQMs) were extracted using yielding a total of 110 IQMs per scan. All IQMs were standardised (z-scored) across scans, and metrics with zero variance were excluded. Principal component analysis (PCA) was applied to the standardised IQM matrix to reduce dimensionality and mitigate collinearity. The smallest number of components explaining at least 95% of the total IQM variance were retained. These PCA-derived IQM components are denoted by $\mathbf{Q}$ and treated as orthogonal summaries of image quality variation.

#### 1.2 Additive Batch-Effect QC Selection

We identified IQM components that predominantly capture additive (mean-shift) batch effects while minimising associations with biological variables or image-derived phenotypes (IDPs). For each IDP $Y$ and each retained IQM component $Q_k$, the following linear mixed-effects models were fitted:

$$Q_k \sim \text{age} + \text{timepoint} + \text{batch} + Y + (1|\text{subject}), \tag{1}$$

$$\text{age} \sim Q_k + \text{timepoint} + \text{batch} + Y + (1|\text{subject}). \tag{2}$$

Equation 1 treats the IQM component as the response variable to directly assess its sensitivity to batch effects after accounting for age, timepoint, and the IDP. Equation 2 serves as a guard model to exclude IQM components that encode age-related biological information. IQM components were retained for additive harmonisation if they were significantly associated with batch ($p < 0.05$) but not significantly associated with age or the IDP ($p > 0.05$).

#### 1.3 Additive Harmonisation Model

For each IDP, additive batch effects were modelled using the selected IQM components within a linear mixed-effects framework:

$$Y = \beta_0 + \boldsymbol{\beta}_Q\mathbf{Q} + \boldsymbol{\beta}_X\mathbf{X} + u + \epsilon, \tag{3}$$

where $Y$ denotes the observed IDP, $\mathbf{Q}$ the matrix of selected IQM components, $\mathbf{X}$ the biological covariates (age and timepoint), $u$ a subject-specific random intercept, and $\epsilon$ residual error. Additive harmonisation was applied by explicitly removing the IQM-associated fixed-effect contribution:

$$Y_{\text{add}} = Y - \boldsymbol{\beta}_Q\mathbf{Q}. \tag{4}$$

#### 1.4 Variance Proxy for Multiplicative Effects

To assess potential multiplicative (scaling) batch effects, a variance proxy was derived from the original IDPs. For each IDP, a biological mixed-effects model excluding batch was first fitted:

$$Y \sim \text{age} + \text{timepoint} + (1|\text{subject}). \tag{5}$$

Let $r$ denote the residuals from this model. A proxy for residual variance was defined as:

$$V = \log(r^2). \tag{6}$$

This variance proxy captures subject-level variability not explained by biological covariates and is used to distinguish site-dependent scaling effects from biologically meaningful heteroskedasticity.

#### 1.5 Multiplicative Batch-Effect QC Selection

To identify IQM components capturing multiplicative batch effects, each IQM component $Q_k$ was modelled as:

$$Q_k \sim \text{age} + \text{timepoint} + \text{batch} + V + (1|\text{subject}), \tag{7}$$

with an additional guard model:

$$\text{age} \sim Q_k + \text{timepoint} + \text{batch} + V + (1|\text{subject}). \tag{8}$$

IQM components were retained for multiplicative correction if they were significantly associated with batch ($p < 0.05$) but not significantly associated with the residual variance proxy or age ($p > 0.05$).

#### 1.6 Multiplicative Harmonisation and Model Comparison

Multiplicative effects are evaluated on additively corrected IDPs in log-space. The multiplicative-stage linear mixed-effects model is written in direct analogy to the additive model (Eq. 3), but with the log-transformed response:

$$\log(Y_{\text{add}}) = \beta_0 + \boldsymbol{\beta}_Q \mathbf{Q} + \boldsymbol{\beta}_X \mathbf{X} + u + \varepsilon. \tag{9}$$

Model comparison is performed between the nested models

$$\log(Y_{\text{add}}) \sim \text{age} + \text{timepoint} + (1|\text{subject}), \tag{10}$$

$$\log(Y_{\text{add}}) \sim \text{age} + \text{timepoint} + \mathbf{Q} + (1|\text{subject}), \tag{11}$$

using a likelihood-ratio test. Multiplicative correction is applied *only* when the full model (Eq. 11) provides a statistically significant improvement in fit over the reduced model (Eq. 10); otherwise, the additively corrected IDP $Y_{\text{add}}$ is retained as the final harmonised output. From the fitted full model (Eq. 9) we extract the QC-associated fixed-effect coefficients $\boldsymbol{\beta}_Q$ (excluding the intercept and covariate coefficients $\boldsymbol{\beta}_X$). The QC-driven linear predictor for observation $i$ is

$$\ell_i = \boldsymbol{\beta}_Q \mathbf{Q}_i. \tag{11a}$$

To obtain a multiplicative factor with unit mean, the linear predictor is centred across observations and exponentiated:

$$M_i = \exp\big(\ell_i - \bar{\ell}\big) = \exp\Big(\boldsymbol{\beta}_Q \mathbf{Q}_i - \frac{1}{N}\sum_{j=1}^{N} \boldsymbol{\beta}_Q \mathbf{Q}_j\Big). \tag{11b}$$

The final harmonised IDP is then given by

$$Y_{\text{harmonised},i} = \frac{Y_{\text{add},i}}{M_i}. \tag{12}$$
